# Long-term air pollution, cardiometabolic multimorbidity, and genetic susceptibility: a multi-state modeling study of 415,855 participants

**DOI:** 10.1101/2022.09.15.22280006

**Authors:** Xu Gao, Meijie Jiang, Ninghao Huang, Xinbiao Guo, Andrea A. Baccarelli, Tao Huang

## Abstract

**Background:** Cardiometabolic multimorbidity (CMM) with at least two cardiometabolic diseases (CMDs) including type II diabetes (T2D), ischemic heart disease (IHD), and stroke, is a global health problem with multiplicative mortality risk and deserves to be investigated as a top priority. Although air pollution is a leading modifiable environmental risk for individual CMD, its impacts on CMM progression were poorly understood.

**Objective:** To elucidate the impact of air pollution on CMM progression, individually and in the context of genetic preposition.

**Design:** Multi-state modeling cohort study.

**Setting:** Data were extracted from the UK Biobank.

**Participants:** 415,855 eligible UK Biobank adults that were free of CMDs at baseline.

**Measurements:** Annual concentrations of particulate matter (PM) with a diameter of ≤2.5 μm (PM_2.5_), 2.5-10 μm (PM_2.5-10_), and ≤10 μm (PM_10_), and nitrogen oxides (NO_x_ and NO_2_) were estimated using Land Use Regression model.

**Results:** During a median follow-up of 8.93 years, 33,375 participants had a first CMD (FCMD), and 3,257 subsequently developed CMM. PM_2.5_, PM_10_, NO_2_, and NO_x_ levels, as well as their combined exposure were associated with increased FCMD risks and even higher risks of CMM. Particularly, per a 5-μg/m^3^ increase in PM_2.5_, risks for FCMD and CMM increased by 27% (95% confidence interval: 20%-34%) and 41% (18%-68%), respectively. By FCMD types, participants with IHD had a higher risk of CMM than those with T2D or stroke. Eighty-five CMD-related genetic variants were associated with CMM trajectories in our study and associations of air pollution with FCMD and CMM risks could be aggravated progressively with increasing genetic risks.

**Limitations:** Other major air pollutants including ozone and SO_2_ were not considered due to the data availability.

**Conclusions:** Air pollution has profound adverse health impacts on the progression of CMM through multi-stage dynamics, especially for individuals with IHD and high genetic risk.

## Introduction

Multimorbidity is defined as the simultaneous presence of two or more chronic diseases and is an increasing global health problem with impaired quality of life, reduced life expectancy, and elevated cost of health care resources by a larger scale than any chronic conditions individually (1). Cardiometabolic multimorbidity (CMM), as one of the most common multimorbidity patterns (2), refers to the coexistence of two or three most prevalent cardiometabolic diseases (CMDs), including type II diabetes (T2D), ischemic heart disease (IHD), and stroke (2, 3). A study based on 91 cohorts identified that any combination of these CMDs was associated with multiplicative mortality risk, and life expectancy was substantially lower in people with CMM with a reduction of life expectancy up to 15 years (3). CMM, therefore, is not simply an accumulation of the three healthy conditions but rather a collision of risk factors promoting death. Up to date, there is a dearth of studies on the determinants of CMM, which is urgently needed to seek appropriate management approaches to early prevent and control CMM development (4).

Ambient air pollution, especially fine particulate matter (PM_2.5_, aerodynamic diameter <2.5 μm), has been recognized as a modifiable risk factor for CMDs (5, 6). For instance, a meta-analysis identified a robust positive association between PM exposure and the incidence of T2D (7). Petal et al. also reported that a 10 μg/m^3^ increase in PM_2.5_ and particulate matter (PM) of sizes 10 μm (PM_10_) was associated with 0.25% higher IHD mortality and 0.27% higher IHD morbidity (8). Short-term air pollution exposure could also increase the daily hospital admissions for ischemic stroke (9). Understanding the impact of air pollution on the dynamics of CMM thereby could hint targeted population of CMM prevention. Nevertheless, this is not sufficient to depict the risks of CMM coming from air pollution exposure without accounting for the temporal development of CMM using traditional time-to-event analyses. Additionally, CMDs have been suggested to be affected mutually by environmental exposures and genetic profiles (10), but knowledge on whether the genetic predisposition of CMM and air pollution could jointly promote the CMM development remains unknown.

Therefore, we aimed to investigate the associations of five pollutants (PM_2.5_, PM_2.5-10_, PM_10_, NO_2_, and NOx) with the CMM trajectories using the scheme of multi-state modeling in the UK Biobank. The multi-state model could evaluate the risk of five transient states (baseline CMD-free to first CMD [FCMD], baseline to death, FCMD to CMM, FCMD to death, and CMM to death) on occupying another state (11). Leveraging in-depth genetic information, this study additionally allowed us to explore the potential synergetic effect of genetic profile and air pollution on CMM.

## Methods

### Study design and participants

UK Biobank is a large population-based prospective cohort study (12). As previously described, about 0.5 million UK residents aged 37-73 years were enrolled from 2006-2010, and the end date of follow-up was 31 December 2018. Their information collected includes lifestyle and health data, physical measurements, and biological samples. After excluding participants without information on air pollution, CMDs, and selected covariates, and those with T2D, IHD, and stroke at baseline, a total of 415,855 participants without diagnosis of the three CMDs were included in our study for further analyses to exclude the impact of baseline CMD on the association between air pollution and CMM trajectories (Figure S1). The UK Biobank was approved by the North West Multicenter Research Ethical Committee. All participants provided informed written consent. This research has been conducted using the UK Biobank Resource under Application Number 44430.

### Cardiometabolic diseases and mortality

Incident IHD, stroke, and T2D cases, as well as relevant mortality, were retrieved from primary care and hospital admission data using the UK National Health Services register. We used the International Classification of Diseases 10th revision (ICD-10) and primary care health records to identify relevant diagnoses. IHD was defined by codes I20 to I25. Stroke was defined by codes I60 to I69. T2D was diagnosed based on the following criteria: having fasting glucose ≥7.0 mmol/L or 2-hour plasma glucose ≥11.1 mmol/L, having physician-diagnosed diabetes, using diabetes medication, or with ICD-10 code E11.

### Exposure assessments

Annual 1-year moving average ambient concentrations of PM_2.5_, PM_2.5–10_, PM_10_, NO_2_, and NO_x_ were calculated with a Land Use Regression (LUR) model. Details about the LUR model are in the supplement methods. Previous reports using leave-one-out cross-validation demonstrated good model performance for PM_2.5_, PM_10_, NO_2_ and NO_x_ (R^2^ of cross-validation =77%, 88%, 87% and 88%, respectively) and a moderate performance for PM_coarse_ (cross-validation R^2^=57%). Concentrations of PM_2.5_, PM_2.5–10_, and NO_x_ were available in 2010 only, while concentrations of PM_10_ (2007 and 2010) and NO_2_ (2005, 2006, 2007, and 2010) were collected for multiple years. Since the baseline enrollment was conducted from 2006 to 2010, to better fit the time frame of baseline survey, we used averaged levels of PM_10_ and NO_2_ in this study.

To explore the combined impact of air pollutants, we weighted the levels of PM_2.5_, PM_10_, NO_x_, and NO_2_ using the β coefficients retrieved from the transition process from baseline to FCMD based on Cox regression models adjusting for all covariates described below. PM_2.5-10_ was removed due to its null associations with CMDs (see the Result section for details). This approach was validated in a 10-fold cross-validation analysis in a previous air pollution-related study based on UK Biobank (13, 14). Weighted indicator (termed as “co-exposure score”) was estimated as: co-exposure score = (β_[PM2.5]_ ×PM_2.5_ + β_[PM10]_ × PM_10_ + β_[NO2]_ × NO_2_ +β_[NOx]_ × NO_x_)×(4/sum of the β coefficients). The co-exposure score ranged from 36.50 to 141.78. Participants were divided into five groups based on the quintiles.

### Genetic information and genetic risk scores

Detailed information about genotyping, imputation, and quality control in UK Biobank has been described previously (15) and provide in supplement methods. Based on previous genome-wide association studies (GWASs) (16-18), we retrieved 74 single-nucleotide polymorphisms (SNPs) for T2D, 55 for IHD, and 32 for stroke (Table S1). We first coded each SNP as 0 and 1 according to the existence of risk alleles to test their independent associations with CMM using MSM and then selected 85 SNPs that were nominal significantly related to any processes of CMM development (see the Result section for details) to create a weighted genetic risk score (GRS). Each selected SNP was then recoded as 0, 1, or 2 according to the number of risk alleles, and multiplied by the logarithmic odds ratios (ORs) obtained from the previous GWAS to calculate the GRS as previously described (19). The GRS ranged from 57.4 to 117.9, with a higher score indicating a higher genetic risk of CMM.

### Covariates

As previously reported (20), the following potential covariates were retrieved from the baseline questionnaires and physical examinations, including age, sex (male/female), body mass index (BMI), years of education (<10 years/ ≥10 years), race (based on UK Biobank question “self-reported ethnic group”, categorized into White, Black, Asian, and other; other included those reported “white and black mixed” and “other ethnic group” to the question), smoking status (never, former, current), employment status (employed/unemployed), total household income (≥£31000/ <£31000), moderate alcohol intake status (yes/no), and high-level physical activity status (yes/no). Detailed measures of the covariates were described in supplement methods.

### Statistical analysis

Survival time for each participant was calculated as the duration from the response date of baseline to the date of incident, death, or date of censoring, whichever came first. We first used Cox regression to examine the associations of five pollutants with FCMD, CMM, and death, separately. Proportional hazard assumptions were not violated based on the Schoenfeld residuals test. Previously described covariates were adjusted in the models. We then used the unidirectional multi-state model (MSM) to assess five pollutants in the temporal disease development from free of CMDs to FCMD, CMM, and death. The MSM was carried out with Markov proportional hazards (21, 22), which is an extension of competing risks survival analysis allowing estimation of the role of a certain factor in different phases of a process. If a participant was recorded with at least two events (CMDs and/or death) at the same time point simultaneously, we theoretically assumed that the order of disease occurrence was T2D, IHD, and stroke according to previous evidence (3, 23, 24), and died eventually. Times for the previous disease were accordingly minus 0.5 days.

First, five transition processes were constructed according to the natural history of CMM and previous studies (24, 25) (transition pattern A, Figure 1): (I) from baseline to FCMD; (II) from baseline to death from a disease other than T2D, IHD, and stroke; (III) from FCMD to CMM; (IV) from FCMD to death from any causes; and (V) from CMM to death from any causes. Processes I, III, and V were the primary ones with the most interest of study in our analysis. Then, to explore which CMD was associated with higher risks of CMM, we further used the MSMs to analyze the effects of air pollutants on different pathways from baseline to CMM by dividing the FCMDs into T2D, IHD, and stroke separately. Thus, along with processes II and V of pattern A, the processes I, III, and IV of pattern A were further divided into nine sub-transitions (Ia-Ic; IIIa-IIIc, IVa-IVc) in pattern B (transition pattern B, Figure 1). Restricted cubic spline regression models were then used to evaluate the dose–response relationship between air pollutants and each CMM processes with 3 knots (10^th^, 50^th^, and 90^th^ percentiles of exposures).

**Figure 1.**
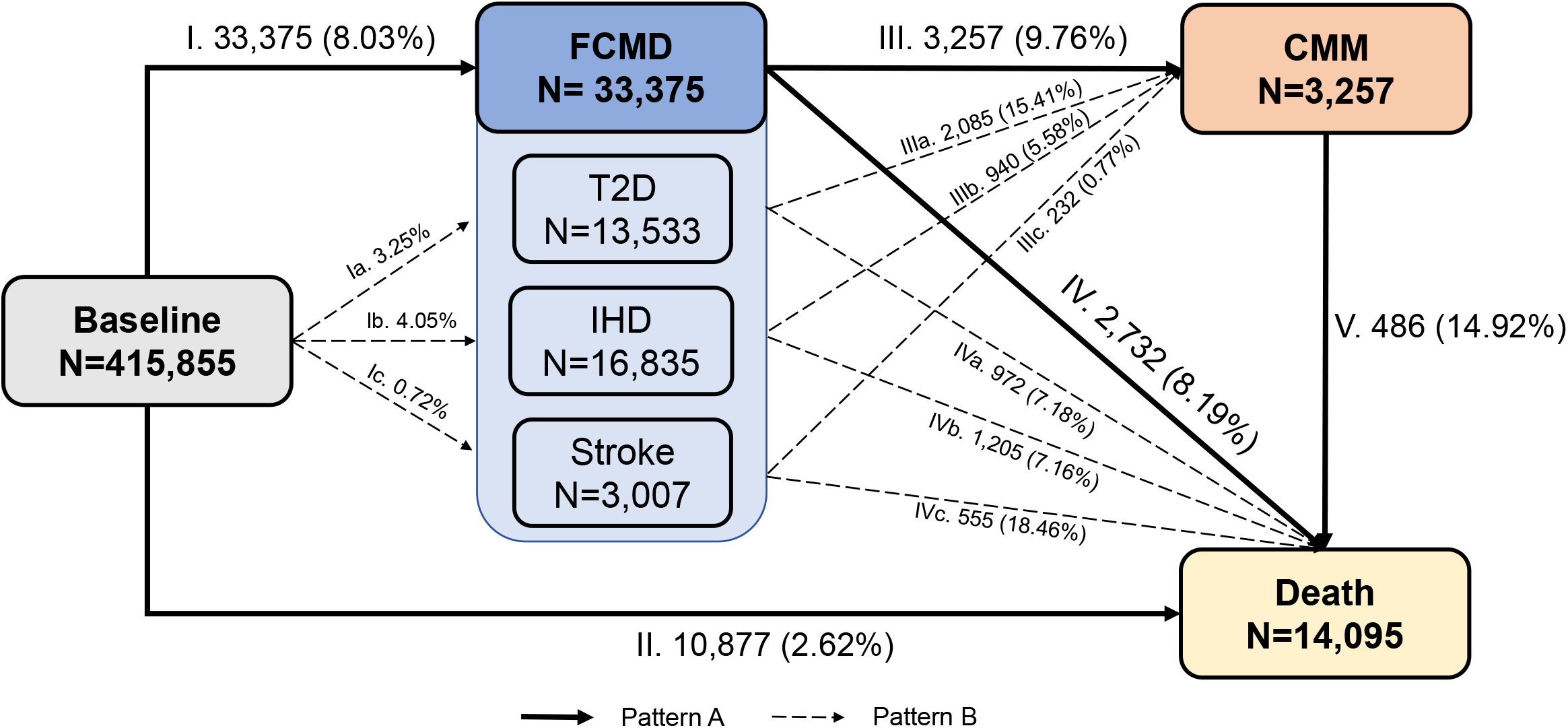
Numbers (percentages) of participants in transition pattern A (solid line) and B (dashed line) from baseline to first cardiometabolic disease (FCMD), cardiometabolic multimorbidity (CMM), and death.

To clarify the effects of genetic susceptibility and air pollution on CMM, we tested the gene–air pollution interaction first using the MSM with an interaction term of the continuous co-exposure score and CMM-related SNPs and continuous GRS to examine the potential modifying effect of genetic background. In the case that interaction effect did not meet criteria for statistical significance (p-values<0.05), we generated a series of categorical variable based on the quintiles of co-exposure score and binary GRS (by median) to assess their joint effects on CMM development.

Six sensitivity analyses were conducted to validate the robustness of our findings. First, we constructed another weighted co-exposure score based on PM_2.5_, NO_2_, and NO_x_ to understand whether the marginal association of PM_10_ with CMM may distort the findings towards null. Then we conducted the MSM of pattern A by adjusting for baseline diet behavior, and levels of low-density lipoprotein, high-density lipoprotein, triglycerides, total cholesterol, systolic blood pressure, and diastolic blood pressure, all of which were established to be related to CMD and their subsequent outcomes (26). Furthermore, we conducted the MSM of pattern A by adjusting for ten genetic principal components of UK Biobank and also in the white participants only to ensure the reliability of our findings across different ethnicities. To understand the impact of mobilization on our findings, we additionally conducted the MSM of pattern A in participants who self-reported that he/she had been living at the same baseline address for more than five years. Last, we evaluated whether the age at FCMD onset (<65 years or ≥65 years) could modify the association between air pollution and subsequent CMM.

All analyses were conducted using R (version 4.0.1). MSM was performed using “mstate” package of R. A two-tailed *p*-value <0.05 indicated statistical significance.

## Results

### Descriptive analysis

Detailed characteristics of the eligible 415,855 participants are shown in Table S2. The mean ± standard deviation (SD) age at baseline was 56.23±8.09 years. Most participants (94.83%) were white. Nearly half of the participants were former or never smokers with moderate alcohol intake, and had a total household income ≥£31,000. During a median follow-up of 8.93 years, a total of 33,375 participants (8.03%) experienced FCMD. Among them, about half experienced IHD (N=16,385), 13,533 experienced T2D, and 3,007 experienced strokes (Figure 1). The mean (SD) ambient levels of PM_2.5_, PM_2.5–10_, PM_10_, NO_2_, and NO_x_ were 9.97 (1.05), 6.42 (0.90), 19.28 (1.95), 29.08 (9.21), and 43.74 (15.56) μg/m^3^, respectively. The pollutants were mutually correlated with each other (Figure S2, *p*-values <0.001).

### Associations between air pollution and cardiometabolic multimorbidity

Patten A was our primary model of CMM development (Figure 1). Fully-adjusted Cox models showed that PM_2.5_, PM_10_, NO_2_, and NO_x_ were associated with the risks of FCMD, CMM, and death, respectively (Table S3). The MSM was therefore useful to uncover the underlying risks of CMM development. Figure 2 depicted that PM_2.5_, NO_2_, and NO_x_ were significantly associated with all CMM processes except for CMM-death. PM_10_ was marginally related to baseline-FCMD and subsequently to CMM. The risk estimates of the four air pollutants were much higher for FCMD-CMM or FCMD-death than for baseline-FCMD. For instance, per 5-μg/m^3^ increment, PM_2.5_ was associated with 27% higher risk of the whole population to develop FCMD from baseline (95% confidence intervals [CIs]: 20%-34%), 41% higher risk of individuals with FCMD to progress from FCMD to CMM (95% CI: 18%-68%), and 35% higher risk of individuals with FCMD to progress from FCMD to death (95% CI: 11%-63%). Same increases in NO_x_ and NO_2_ demonstrated similar significant magnitudes but with reduced estimates. PM_2.5-10_ had null associations with all CMM processes. Monotonic increasing dose-response relationships were observed for the associations of PM_2.5_, NO_x_, and NO_2_ with the processes of baseline-FCMD, FCMD-CMM, and FCMD-death (Figures 3 & S3).

**Figure 2.**
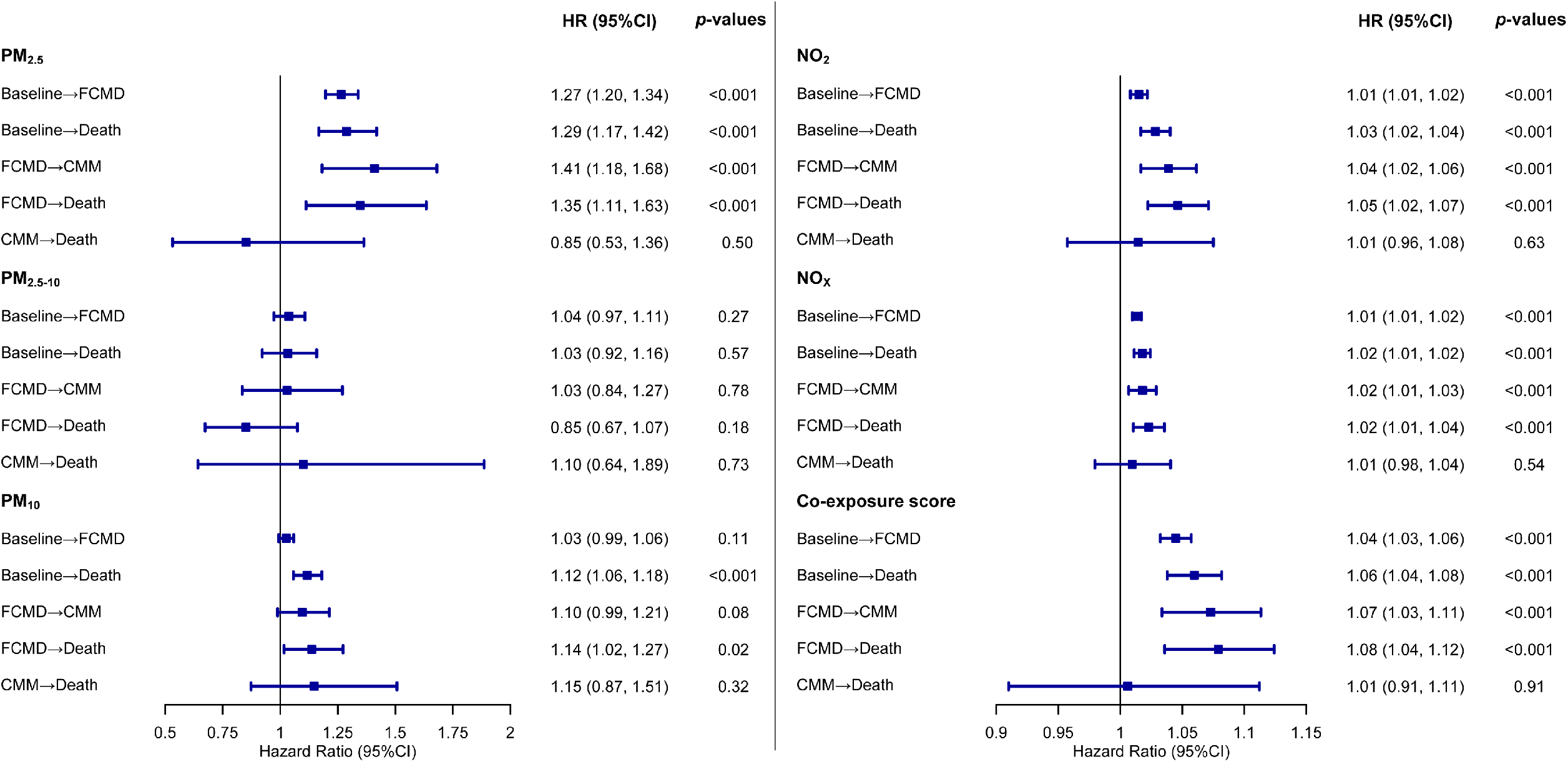
Associations of five air pollutants and co-exposure score with the risks of first cardiometabolic disease and cardiometabolic multimorbidity of pattern A using multi-state model Models adjusted for age, sex, ethnicity, BMI, years of education, smoking status, moderate alcohol intake, high-level physical activity, total household income, and employment status. Estimates of air pollutants were demonstrated per 5-μg/m3 increase and estimates of co-exposure score were demonstrated per one SD increase. Dots: Point estimate; Error bar: 95% confidence limits;

**Figure 3.**
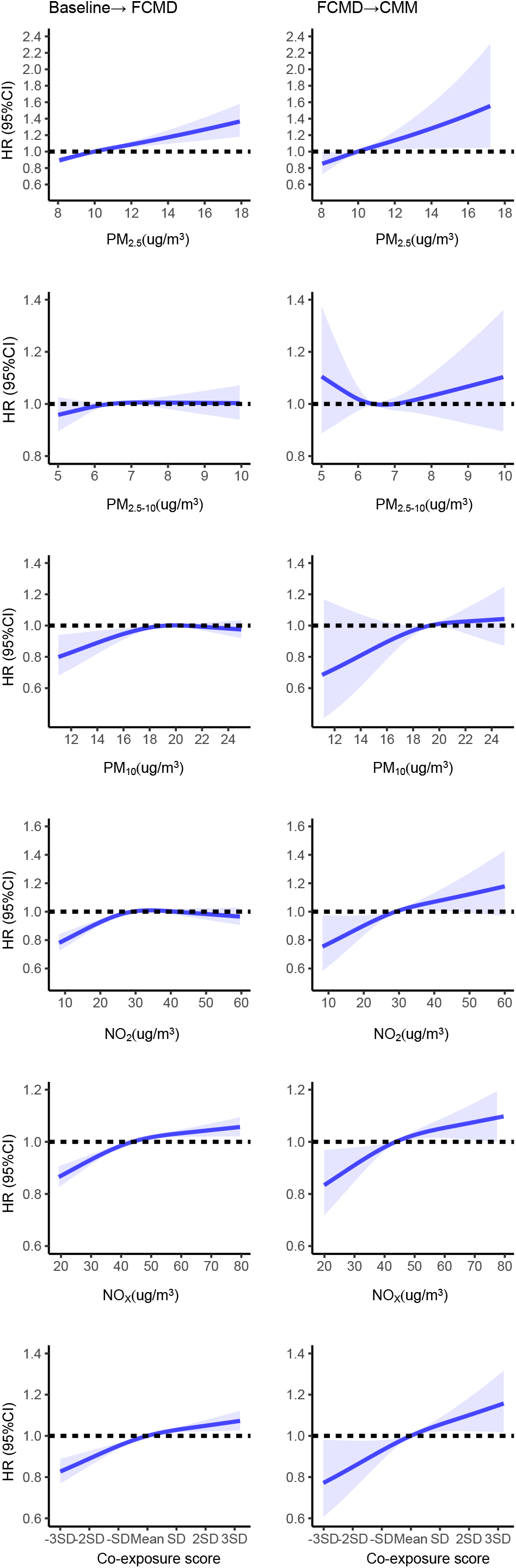
Graphs of the dose-response relationships of five air pollutants and co-exposure score with the processes from baseline to FCMD and from FCMD to CMM of pattern A Solid lines: point estimates; Shadows: 95% confidence limits;

Strong associations between the co-exposure score based on PM_2.5_, PM_10_, NO_x_, and NO_2_ and the CMM processes were also observed, except for CMM-death (Figures 2 & S3-S4). Compared to the 1^st^ quintile of the co-exposure score, the 5^th^ quintile was associated with 14% (95% CI: 10%-18%), 22% (95% CI: 7%-38%), and 22% (95% CI: 7%-40%) higher risks for processes of baseline-FCMD, FCMD-CMM, and FCMD-death, respectively. A sensitivity analysis using the modified co-exposure score based on PM_2.5_, NO_2_, and NO_x_ also yielded similar estimates (Table S4).

In transition pattern B which classified FCMDs into three CMDs (Figure 1), we found baseline participants were more likely to initially develop T2D than IHD or stroke (Tables S5 & S6). However, FCMD participants with IHD were more likely to develop CMM than those with T2D or stroke in response to PM_2.5_, NO_2_, NO_x_, and the co-exposure score (Table 1). For instance, per a 5-μg/m^3^ increase in PM_2.5_, the corresponding hazard ratios (HRs) of CMM were 1.51 (95% CI: 1.08-2.10), 1.05 (95% CI: 1.01-1.10), and 1.01 (95% CI: 0.88-1.15) for FCMD participants with IHD, T2D, and stroke, respectively.

**Table 1.** Associations of five air pollutants and co-exposure score with the risks of first cardiometabolic disease, cardiometabolic multimorbidity, and death after the first cardiometabolic disease of pattern B using multi-state model ^a^

| Air pollutants | PM<sub>2.5</sub> | PM<sub>2.5-10</sub> | PM<sub>10</sub> | NO<sub>2</sub> | NO<sub>x</sub> | Co-exposure score<sup>a</sup> |
| --- | --- | --- | --- | --- | --- | --- |
| Trajectories | HR (95% CI) | HR (95% CI) | HR (95% CI) | HR (95% CI) | HR (95% CI) | HR (95% CI) |
| **Baseline→T2D** | **1.47 (1.34, 1.60)** | 1.86 (1.12, 3.08) | **1.79 (1.40, 2.30)** | **1.21 (1.15, 1.28)** | **1.12 (1.09, 1.15)** | **1.08 (1.06, 1.10)** |
| **Baseline→IHD** | **1.13 (1.04, 1.22)** | 0.76 (0.48, 1.21) | 0.76 (0.61, 0.95) | 0.98 (0.93, 1.02) | **1.03 (1.00, 1.05)** | 1.01 (1.00, 1.03) |
| **Baseline→Stroke** | **1.23 (1.02, 1.48)** | 1.89 (0.65, 5.51) | 1.26 (0.75, 2.11) | 1.04 (0.93, 1.16) | **1.05 (0.98, 1.11)** | 1.03 (0.99, 1.08) |
| **T2D→CMM** | **1.05 (1.01, 1.10)** | 1.01 (0.96, 1.06) | 1.01 (0.99, 1.04) | **1.00 (1.00, 1.01)** | **1.00 (1.00, 1.01)** | 1.05 (1.00, 1.10) |
| **IHD→CMM** | **1.51 (1.08, 2.10)** | 0.90 (0.12, 6.56) | 1.24 (0.46, 3.35) | **1.28 (1.04, 1.58)** | **1.15 (1.04, 1.27)** | **1.10 (1.02, 1.18)** |
| **Stroke→CMM** | 1.01 (0.88, 1.15) | 0.92 (0.78, 1.09) | 0.98 (0.91, 1.06) | **1.01 (1.00, 1.02)** | 1.00 (0.98, 1.01) | 0.99 (0.85, 1.14) |
| **T2D→Death** | 1.03 (0.97, 1.10) | 0.99 (0.92, 1.07) | 1.01 (0.98, 1.05) | 1.00 (1.00, 1.01) | 1.00 (1.00, 1.01) | 1.04 (0.97, 1.11) |
| **IHD→Death** | **1.10 (1.04, 1.17)** | 0.95 (0.88, 1.02) | **1.03 (1.00, 1.07)** | **1.01 (1.01, 1.02)** | **1.01 (1.00, 1.01)** | **1.13 (1.06, 1.20)** |
| **Stroke→Death** | 1.01 (0.92, 1.10) | 0.94 (0.84, 1.04) | 1.01 (0.96, 1.07) | 1.01 (1.00, 1.02) | 1.00 (0.99, 1.01) | 1.02 (0.93, 1.12) |

a: Models adjusted for age, sex, ethnicity, BMI, years of education, smoking status, moderate alcohol intake, high-level physical activity, total household income, and employment status. Estimates of air pollutants were demonstrated per 5-μg/m<sup>3</sup> increase and estimates of co-exposure score were demonstrated per one SD increase. Bolded-values were statistically significant (*p*-values <0.05).

### Genetic susceptibility of cardiometabolic multimorbidity and joint effects with air pollution

For the retrieved CMD-related SNPs, 38 T2D-related, 32 IHD-related, and 15 stroke-related loci were nominally associated with any CMM trajectories of pattern A (Tables S1 & S7 a-c, nominal *p*-values <0.05). Genes *ABO, EDNRA, FURIN, MIA3*, and *CDKN2B-AS1* had ≥2 unique SNPs, and *CDKN2B-AS1* had the most including rs3217992, rs4977574, and rs7859727. The weighted GRS based on these CMM-related loci was robustly associated with CMM transitions (Table S7). However, none of the SNPs or the continuous GRS could interact with the co-exposure score in relation to CMM trajectories (Table S6 d-f, interaction FDR-corrected *p*>0.05; Table S9, interaction *p*-values >0.1). And in the mutual adjustment model, the coefficients of GRS and the co-exposure score remained essentially unchanged compared to models with either factor (Table S9). We suggested that the environmental and genetic factors were independently associated with CMM development.

Given no interaction was observed, we generated a series of categorical variable based on the quintiles of co-exposure score and dichotomized GRS (by median) to test the joint effects of both factors. We observed gradients in associations of higher GRS and co-exposure score for the processes of baseline-FCMD and FCMD-CMM (Figure 4). For instance, FCMD participants with the 5^th^ quintile co-exposure score and a high GRS had a higher risk of CMM compared to those with the 1^st^ quintile of co-exposure score and a low GRS with a HR of 1.40 (95% CI: 1.16-1.70) (Table S19). But no significant joint effects on baseline-death, FCMD-death, or CMM-death was found.

**Figure 4.**
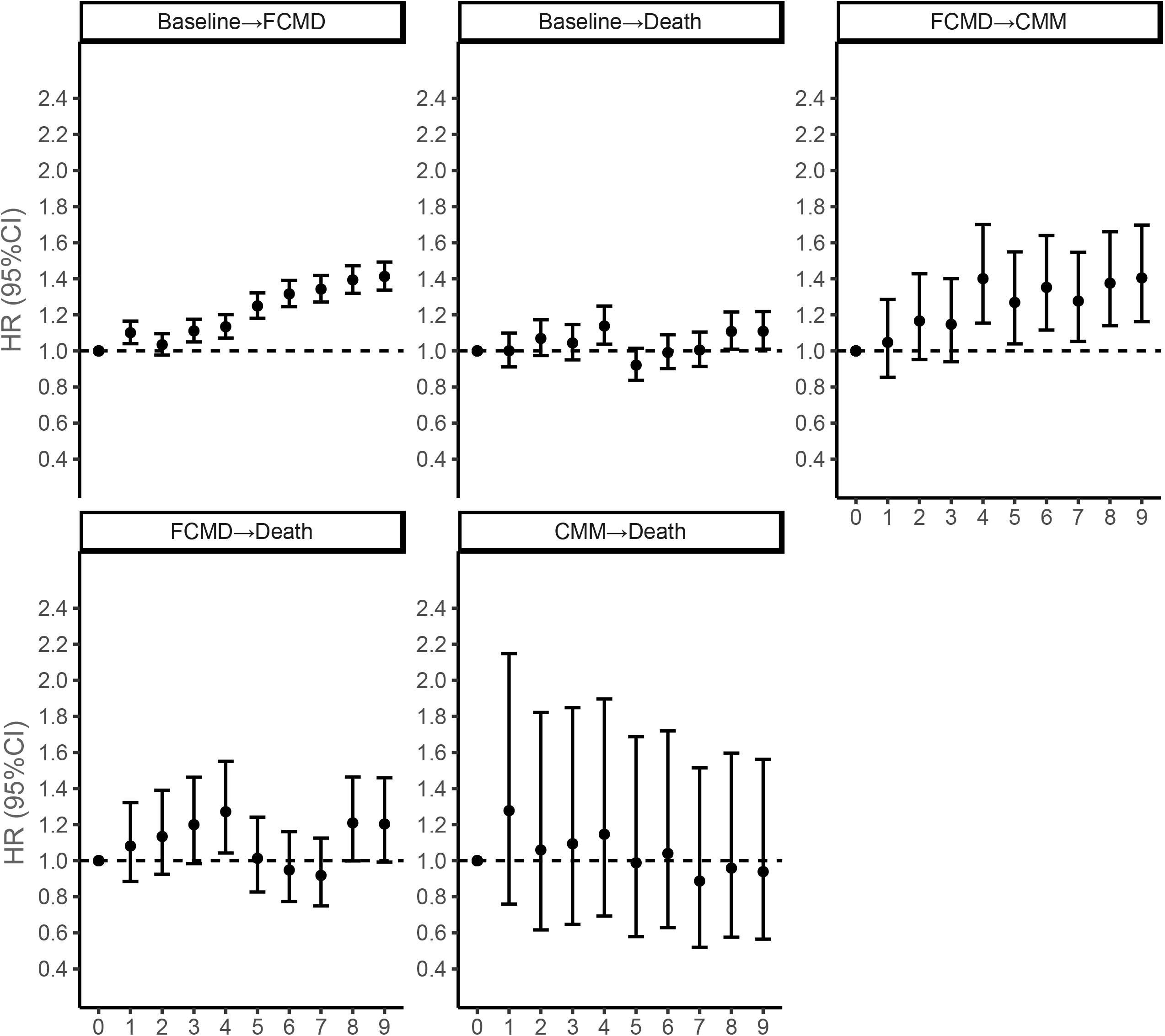
Joint associations of weighted genetic risk and air pollution levels of five transition processes on pattern A using multi-state model 0: low GRS & 1^st^ quintile of co-exposure score (reference); 1: low GRS & 2^nd^ quintile of co-exposure score; 2: low GRS & 3^rd^ quintile of co-exposure score; 3: low GRS & 4^th^ quintile of co-exposure score; 4: low GRS & 5^th^ quintile of co-exposure score; 5: high GRS & 1^st^ quintile of co-exposure score; 6: high GRS & 2^nd^ quintile of co-exposure score; 7: high GRS & 3^rd^ quintile of co-exposure score; 8: high GRS & 4^th^ quintile of co-exposure score; 9: high GRS & 5^th^ quintile of co-exposure score;

### Sensitivity analyses

In the model which further adjusted for baseline diet behavior, lipoprotein levels, and blood pressure, (Table S11), the estimates of PM_2.5_, NO_2_, NO_x_, and the co-exposure score remained essentially unchanged in relation to the risks of CMM development. Results from models with additional adjustment with genetic principal components (Table S12) and from models conducted in white participants only (Table S13) also showed similar risks, both of which suggested the robustness of our findings across the ethnicities. Similar estimates were also yielded in participants living in the address for more than five years (Tables S14), suggesting that out primary results were fairly robust regardless of the mobilization. Moreover, although the interactions between the age of FCMD onset (<65 years or ≥65 years) and air pollution on the CMM risks were not significant (Table S15, interaction *p*-values >0.2), air pollution–CMM associations were stronger among the elderly aged <65 years than those aged ≥65 years.

## Discussion

Utilizing the prospective data of ∼0.42 million participants of the UK Biobank, we uniquely found that PM_2.5_, NO_2_, and NO_x_ levels were associated with the CMM individually and jointly as a weighted co-exposure score in a multi-state nature. For participants with any FCMD, those with IHD had higher risks of incident CMM and death than those who had T2D or stroke as FCMD. Risks for developing FCMD and CMM under long-term air pollution could be enhanced by the CMM genetic susceptibility. Our findings will help broaden the scope of public health recommendations encompassing patients with CMDs and to better characterize and distinguish individuals with a high risk of developing CMM under environmental insults.

To date, there is an evident lack of studies that have specifically investigated the temporal association between air pollution and CMM through methods that involve a stochastic process covering the possible stages of multimorbidity rather than simply counting the numbers of non-communicable diseases without considering disease-disease interactions (27). This study is the first investigation showing the critical role of air pollution on the dynamics of CMM, which indicates that historical air pollution exposure, especially to PM_2.5_ may have profound and lasting impacts on cardiovascular and metabolic systems. In consistent with previous studies of individual CMDs (6, 7, 28), our study yielded robust associations of air pollution, especially PM_2.5_, NO_2_, and NO_x_, with FCMD. Moreover, we identified a stepwise increasing magnitude of the associations of each air pollutant with the risks of CMM and death after the occurrence of FCMD. This pattern highlights the value of the secondary prevention of individuals with FCMD to mitigate the additive risks of subsequent CMDs or death. Additionally, although not statistically significant, individuals with a younger age at FCMD diagnosis (<65 years) had higher CMM risk than those older. Future studies are warranted to elucidate whether the age at FCMD onset could be recognized as a potential risk indicator of subsequent CMM risk for the precision prevention of CMM along with environmental risk factors.

More intriguingly, we found that participants were more likely to develop T2D first from baseline under air pollution. This may be plausible that increased blood glucose level has been recognized as a critical pathophysiology mechanism for the risks of IHD and stroke (23). We thus believed that T2D could occur ahead of the onset of IHD or stroke and could be easily diagnosed in routine health examinations. Furthermore, previous evidence implied that those with IHD or stroke could have relatively higher glucose levels or were pre-diabetic because cardiovascular events may impair fasting glucose (29). Even not yet reached the diagnosis criteria of T2D, we anticipated those with IHD or stroke to be more likely to develop T2D and CMM. However, in our study, participants with IHD as FCMD showed higher risks of developing CMM or death than others who first had T2D and stroke. The attenuated risks related to stroke could be explained by better medical care of stroke patients than those with IHD because of the sudden onset of stroke and higher mortality than IHD. Additionally, stroke patients usually receive better and systematic blood pressure and glucose management over rehabilitation period which may considerably lower their risks of IHD, T2D, and/or death (30, 31). Therefore, for areas with relatively higher stroke incidence and heavier burden of air pollution, for instance, in developing countries, CMM prevention for stroke patients may be more than valuable (32). Additionally, except for the limited death numbers, null associations between air pollution and the CMM-death process may be explained by better medical care for CMM patients. A recent study of CMM and mortality in UK general practices have demonstrated that compared with people with individual CMDs, those with multimorbidity could have a higher likelihood of receiving evidence-based treatment to lower the exceeded risk of coexisting cardiovascular and metabolic diseases (33).

Accumulating evidence has suggested the biological pathway of CMD resulting from air pollution (34), including pro-hypertension, insulin resistance, and endothelial damage resulting from oxidative stress and/or inflammation, which could further promote CMM (28, 34). Beyond these common etiological pathways, a recent study showed that the comorbidity of cardiovascular and metabolic diseases could be explained by overlapping genetic profiles along with protein-protein interactions (PPIs) as a complex network (35). In line with this, our study found that four out of the five CMM-related genes (i.e., *ABO, EDNRA, FURIN*, and *MIA3*) that had ≥2 unique SNPs were related to protein-coding and could additionally participate in PPIs of the cardiovascular network. Gene *CDKN2B-AS1* with the most CMM-related SNPs is not a protein-coding gene, but it produces a long non-coding RNA that interacts with nearby genes and impairs the interferon-γ signaling response to elevate the susceptibility of CMDs (36). Since this gene has also been related to multiple aging-related diseases including cancer and glaucoma, as well as abnormalities in blood cell counts (37), more work is required to uncover its role in the development of multimorbidity. Moreover, insignificant interaction between air pollution and GRS was observed, but moderate additive effect of genetic background on the associations of air pollution with the baseline-FCMD and FCMD-CMM processes were shown. Therefore, we believe that genetic susceptibility could be considered in CMM prevention in response to air pollution. Such mild clues based on SNPs reported by previous GWAS of CMDs further suggest the identification of novel CMM-related variants is highly recommended to empower the risk prediction of CMM.

This study has several strengths including the large sample size and detailed records of CMD with information on lifestyle and covariates that allowed us to conduct the state-of-art multi-state analysis. Several limitations should also be noted in the result interpretation. First, we only focused on the CMM defined with the co-occurrence of three diseases including IHD, stoke, and T2D only because this multimorbidity pattern is the most observed and studied in previous literature (2, 3). Other important CMDs such as hyperlipidemia, and nonalcoholic fatty liver disease could be investigated in future studies. Second, UK Biobank is a volunteer cohort with participants who were likely healthier than the general population, and we excluded those with pre-existing CMDs. Both may cause selection bias influencing the effect of air pollution on CMM development towards null as relatively healthier people usually have lower risks of CMDs. Furthermore, the air pollution data was mostly a single measurement of the annual average level without any further measures within a shorter time interval. This prohibited further explorations on the lag or short-term (<1 month) influences of air pollution. Meanwhile, we did not have the data on other major air pollutants, such as ozone and sulfur dioxide, which have been associated with CVD and T2D (38, 39) and could be studied in future multi-state investigations. Additionally, in processing the data to match the multi-state algorithm, we theoretically ascertained the occurrence orders of events if subjects have two or more events reported on the same day. This may reduce the power of MSM but very slightly because the number of cases with such conditions was comparatively tiny (N=1192, ∼3.6% of FCMD participants). Moreover, despite that we created the GRS using a GWAS of CMM based on well-established SNPs that were highly related to any CMD in previous GWASs, its generalizability in predicting CMM may be required to be further validated. Last, we did not have the drug use information of the participants during follow-up, which may considerably mitigate the risks of subsequent adverse cardiometabolic health outcomes. Given the CMD information was mostly retrieved from linkage to electronic health records, we believe that most CMD participants received appropriate treatment. Such well-received CMD management may have underestimated the substantial relevant detrimental impacts of air pollution on CMM development we observed.

Notwithstanding the improved awareness of the public on the harms of air pollution, knowledge is quite limited about whether the impact of environmental exposure on cardiovascular or metabolic systems may further lead to relevant comorbidities. In this study, we suggest air pollution could increase the risk of CMM in almost a decade through multi-stage dynamics, especially for FCMD participants with IHD and participants with high genetic risk. Our results further elucidate the importance of CMM prevention after long-term air pollution exposure regardless of the genetic risk, especially for locations where air quality is poorer than in the UK and the burden of CMDs resulting from air pollution is thus expected to be heavier. For health professionals, general practitioners, and stakeholders, more efforts may be warranted to better protect the vulnerable population from environmental risks of CMM.

## Data Availability

Data are available in a public, open access repository. This research has been conducted using the UK Biobank Resource under Application Number 44430. The UK Biobank data are available on application to the UK Biobank (www.ukbiobank.ac.uk/).

## Acknowledgments

We thank Dr. Chen Chen for the language assistance.

## Supplementary Materials

**Figure S1** Flowchart of participant selection

**Figure S2** Correlation matrix of the ambient levels of five air pollutants

**Figure S3** Graphs of the dose-response relationships of five air pollutants and co-exposure score with the trajectories of cardiometabolic multimorbidity of pattern A (a: PM_2.5_; b: PM_2.5–10_; c: PM_10_; d: NO_2_, e: NO_x_, f: co-exposure score)

**Figure S4** Associations of categorical co-exposure score (quintiles) with the trajectories of cardiometabolic multimorbidity of pattern A using multi-state model Models adjusted for age, sex, ethnicity, BMI, years of education, smoking status, moderate alcohol intake, high-level physical activity, total household income, and employment status.

**Table S1** Characteristics of genetic variants associated with type II diabetes, ischemic heart disease, and stroke

**Table S2** Baseline characteristics of 415,855 participants by incident disease status during follow-up

**Table S3** Associations of five air pollutants with the risks of first cardiometabolic disease, cardiometabolic multimorbidity, and mortality using Cox model

**Table S4** Associations of the co-exposure score based on PM_2.5_, NO_2_, and NO_x_ with the trajectories of cardiometabolic multimorbidity of pattern A using multi-state model

**Table S5** Associations of air pollution with the trajectories of cardiometabolic multimorbidity of pattern B using multi-state model

**Table S6** Associations of categorical co-exposure score (quintiles) with the trajectories of cardiometabolic multimorbidity of pattern B using multi-state model

**Table S7** Associations of CMD-related genetic variants with the trajectories of cardiometabolic multimorbidity of pattern A using multi-state model (a-c) and their interactions with co-exposure score (d-f)

**Table S8** Associations of weighted genetic risk score with the trajectories of cardiometabolic multimorbidity of pattern A using multi-state model

**Table S9** Mutual associations of co-exposure score and genetic risk score with the trajectories of cardiometabolic multimorbidity of pattern A using multi-state model in model with and without interaction terms

**Table S10** Joint associations of weighted genetic risk score and co-exposure score of five transition states on pattern A using multi-state model

**Table S11** Associations of air pollution with the trajectories of cardiometabolic multimorbidity of pattern A using multi-state model additionally adjusted for baseline diet behaviors, cholesterol levels, and blood pressure

**Table S12** Associations of air pollution with the trajectories of cardiometabolic multimorbidity of pattern A using multi-state model additionally adjusted for genetic principal components

**Table S13** Associations of air pollution with the trajectories of cardiometabolic multimorbidity of pattern A using multi-state model in white participants

**Table S14** Associations of air pollution with the trajectories of cardiometabolic multimorbidity of pattern A using multi-state model in participants living in the baseline address for more than five years

**Table S15** Associations of air pollution with the trajectories of cardiometabolic multimorbidity of pattern A using multi-state model by the age of having the first cardiometabolic disease

