## Supplementary figures and images for "Long-term air pollution, cardiometabolic multimorbidity, and genetic susceptibility: a multi-state modeling study of 415,855 participants"

### Figure S1

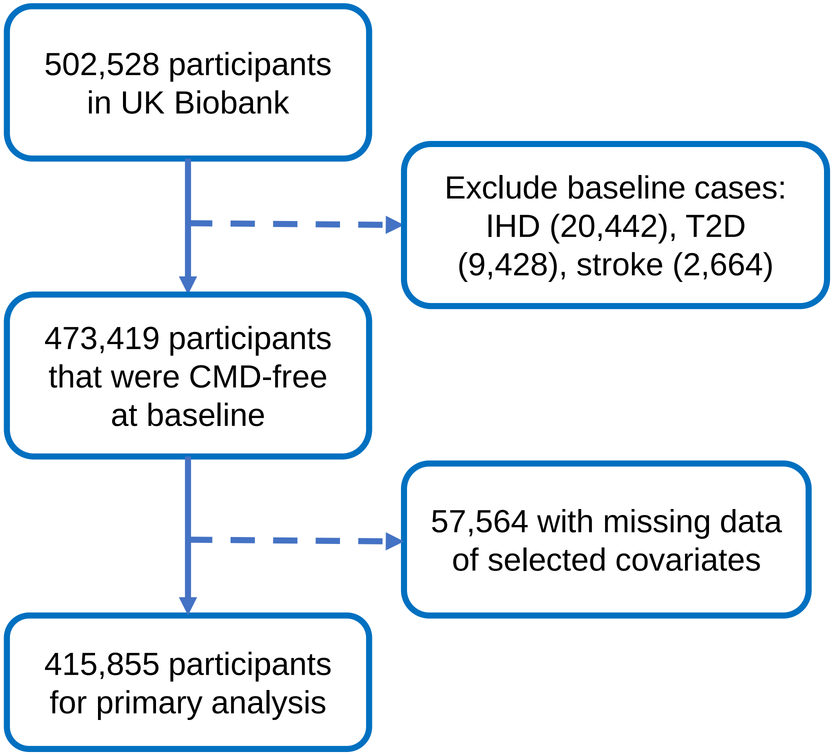

### Figure S2

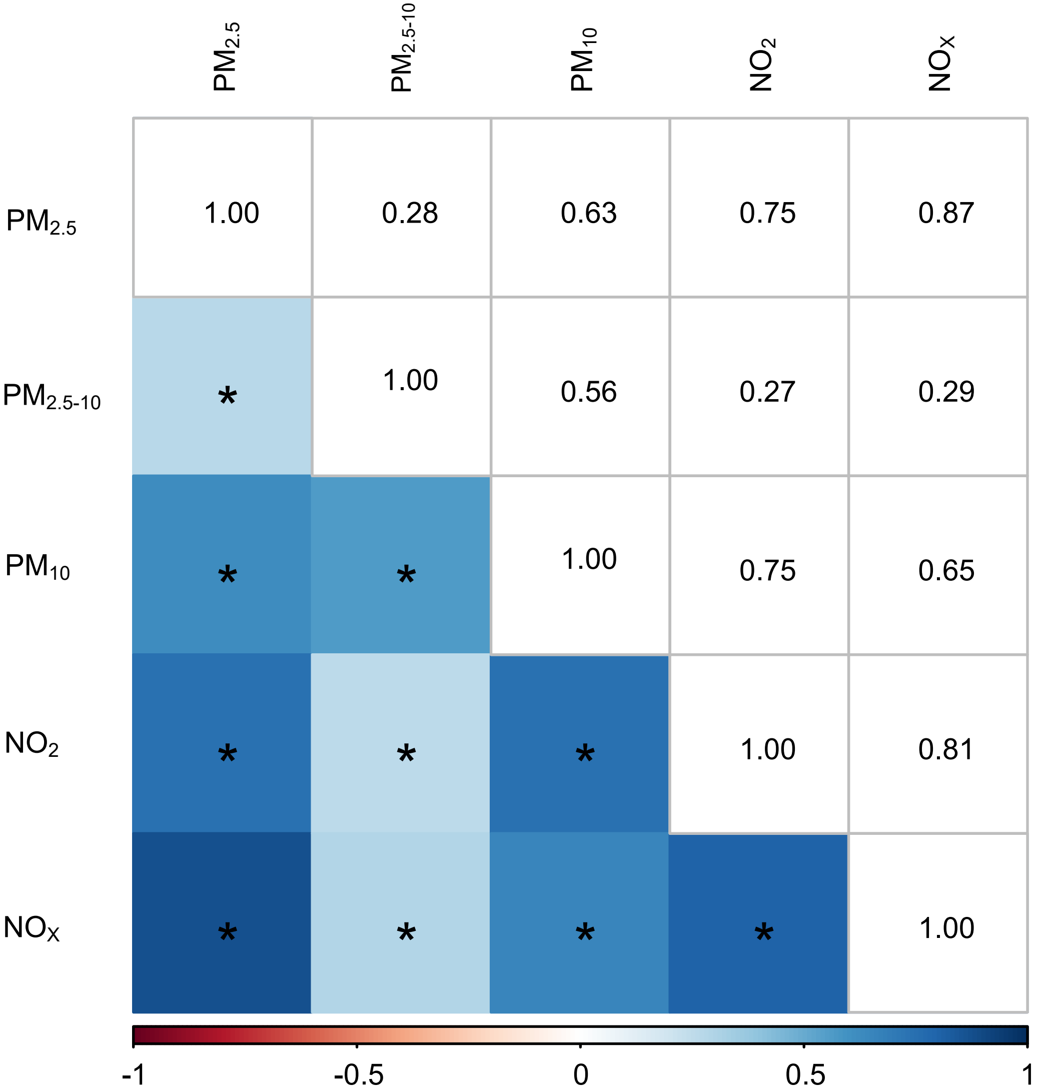

### Figure S3

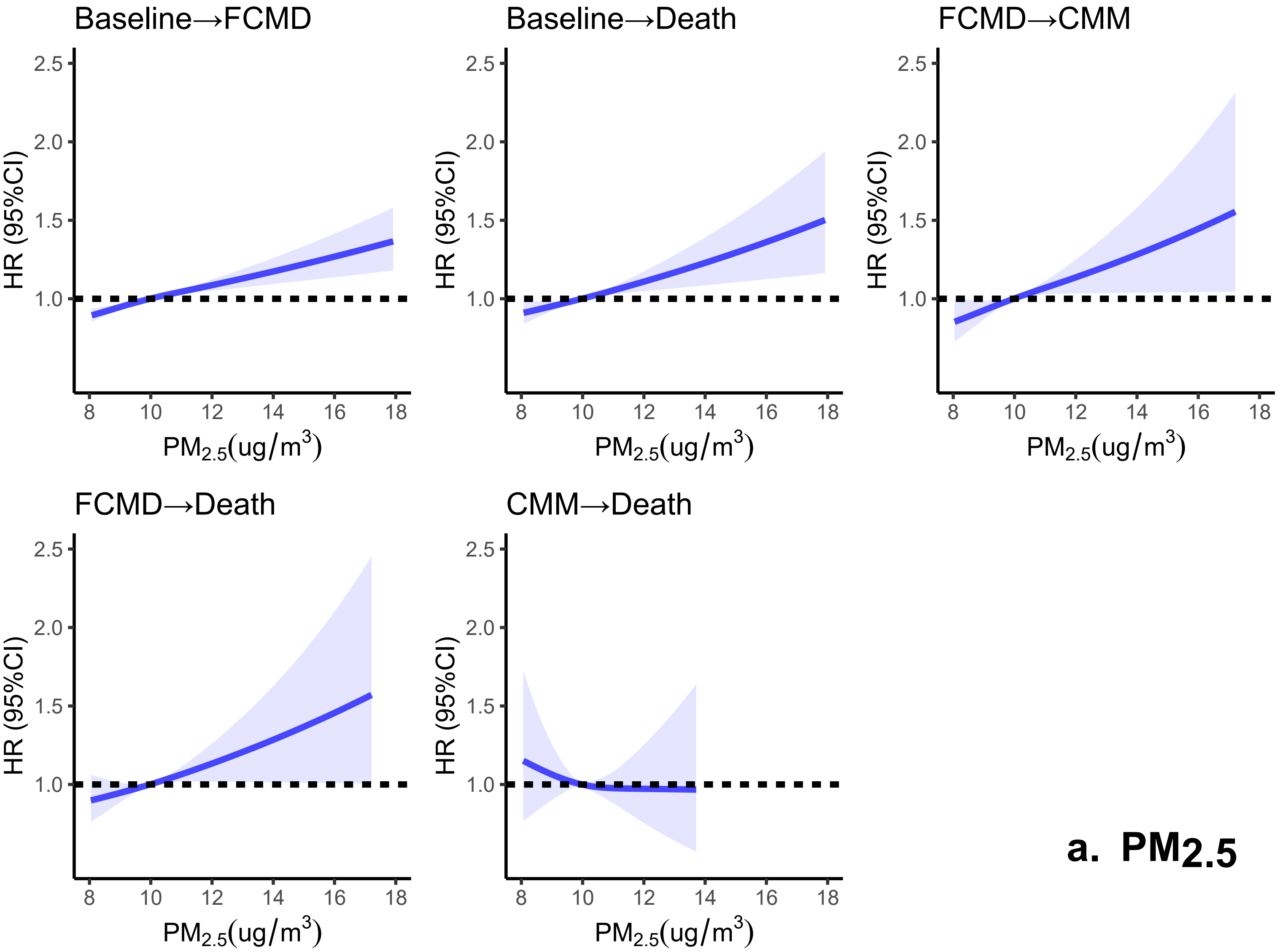

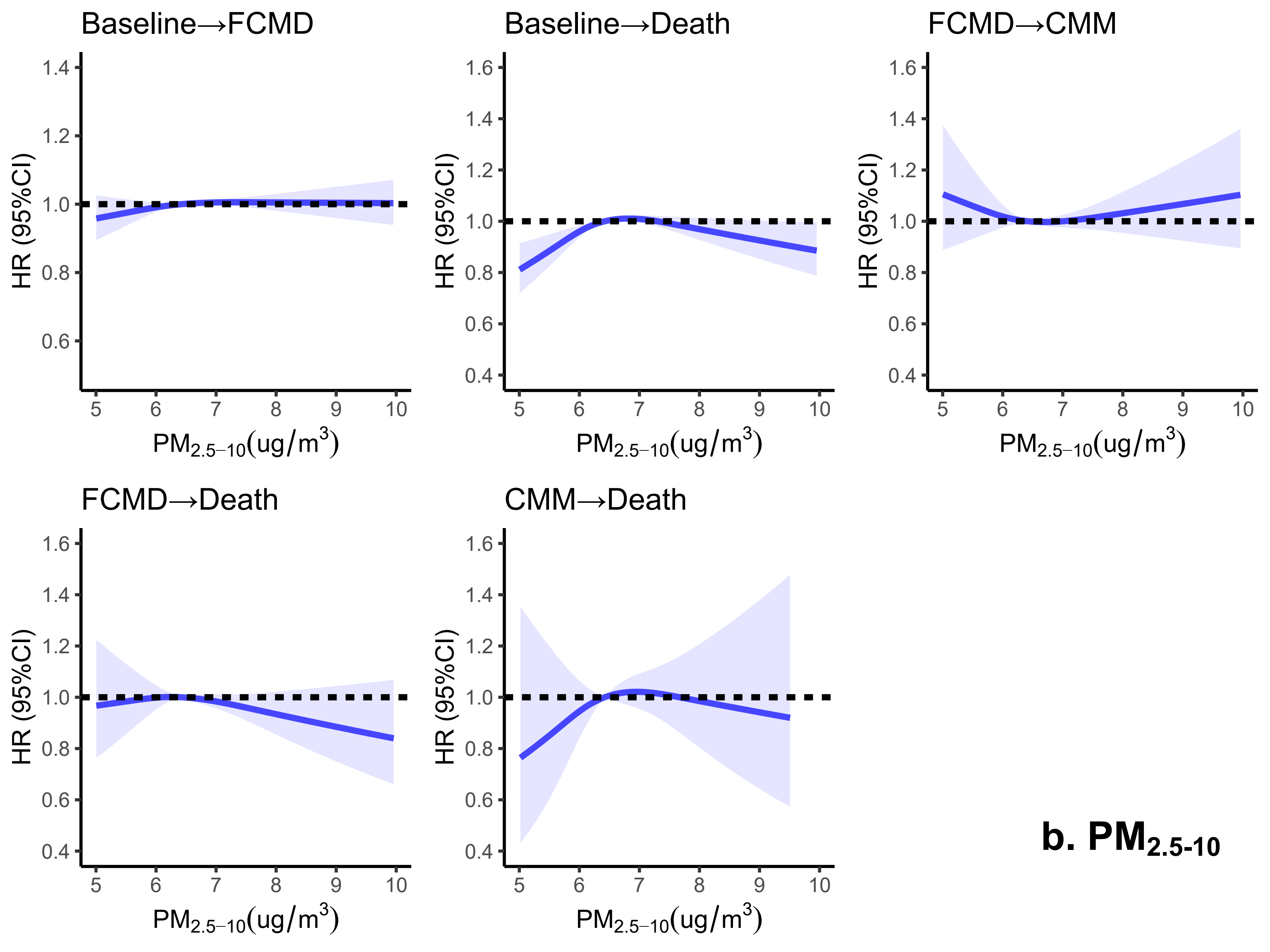

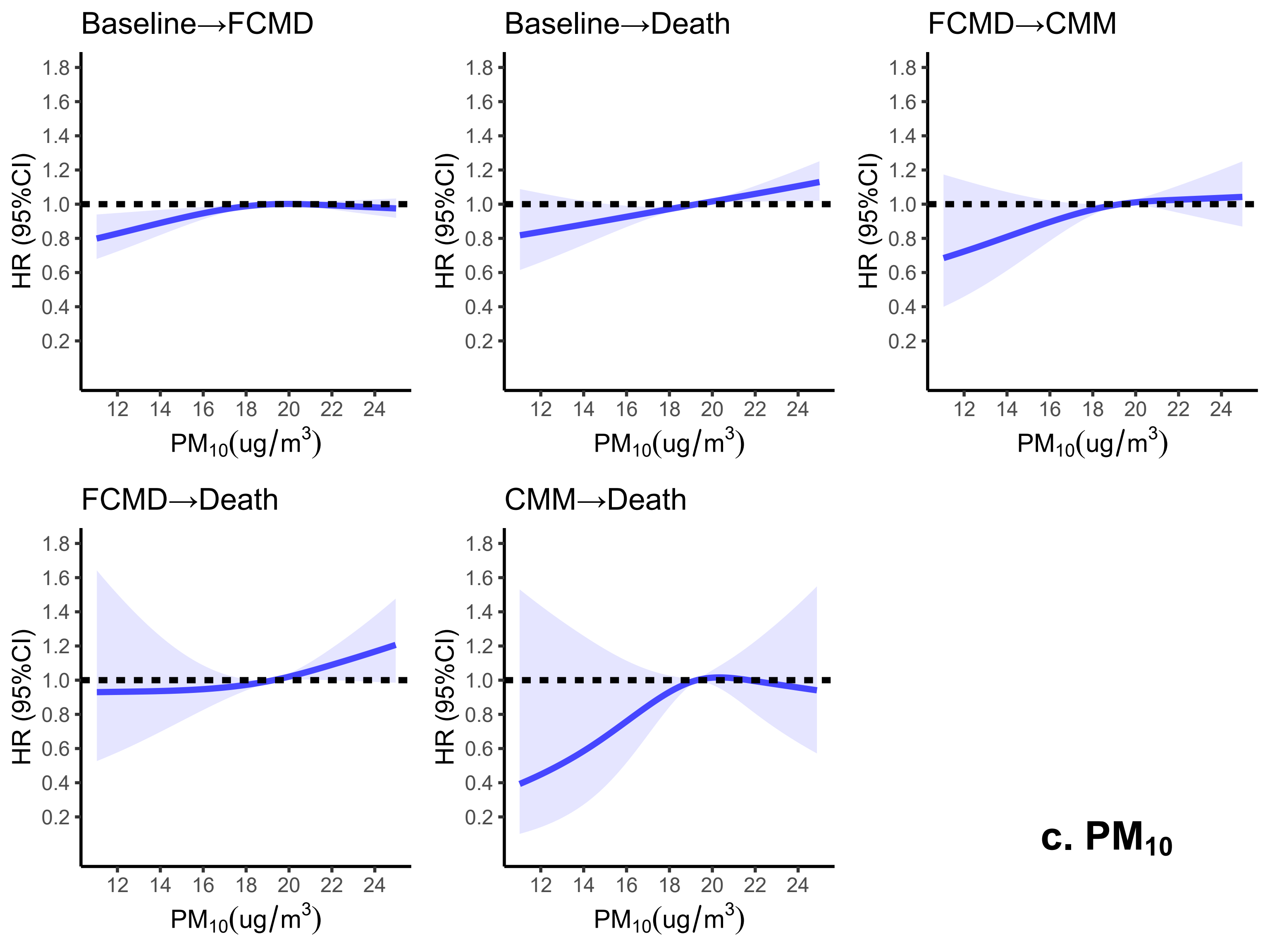

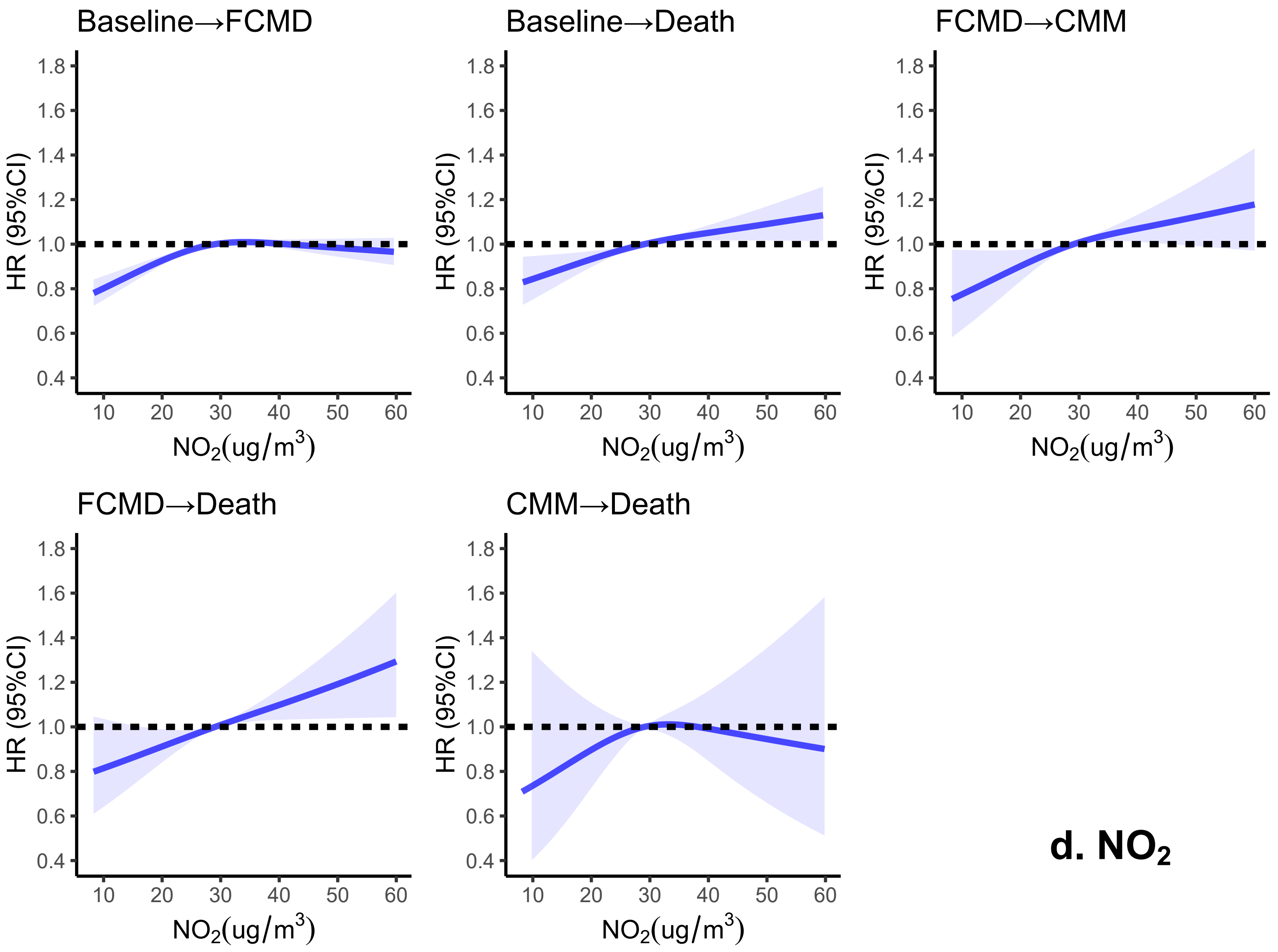

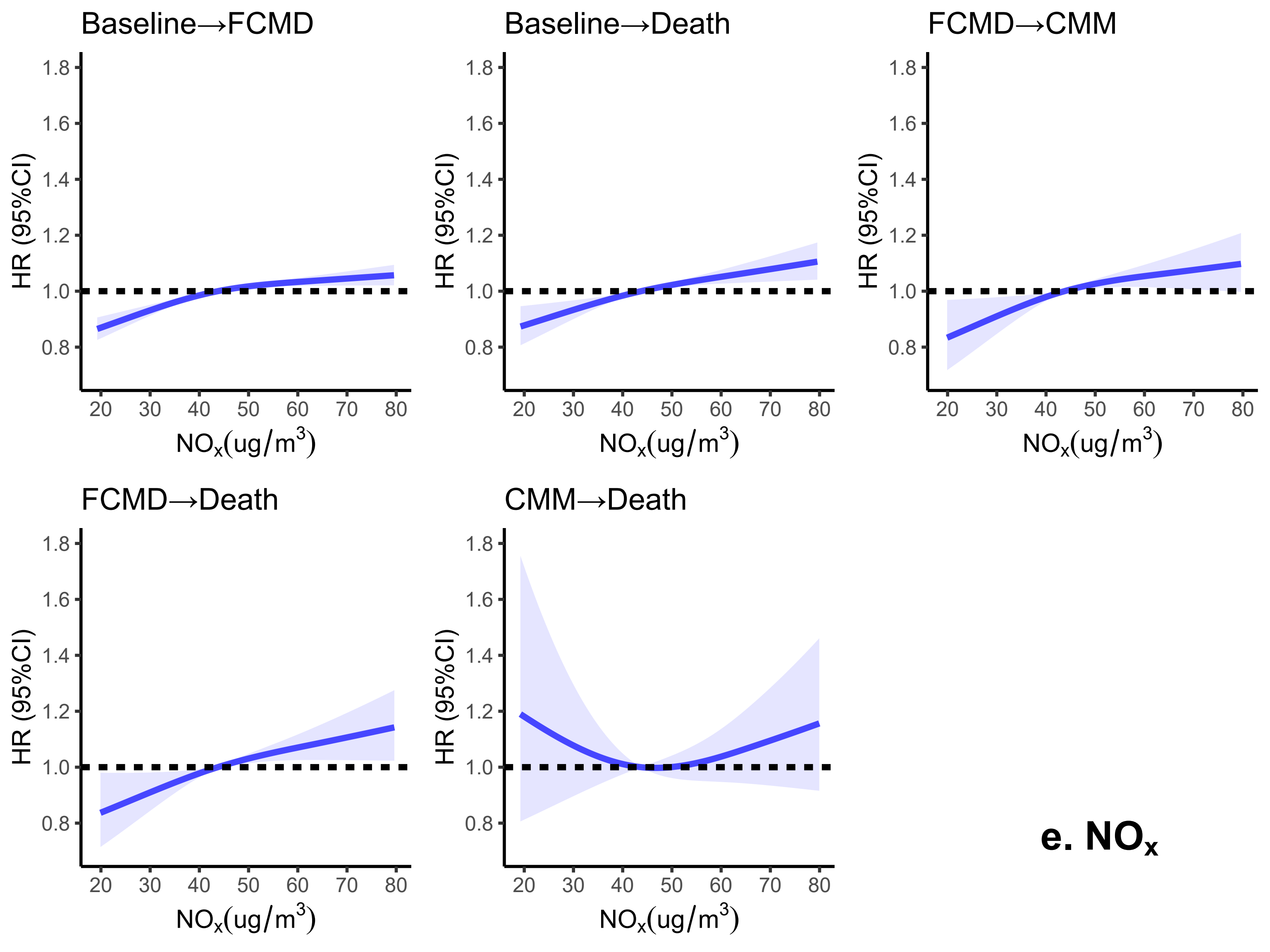

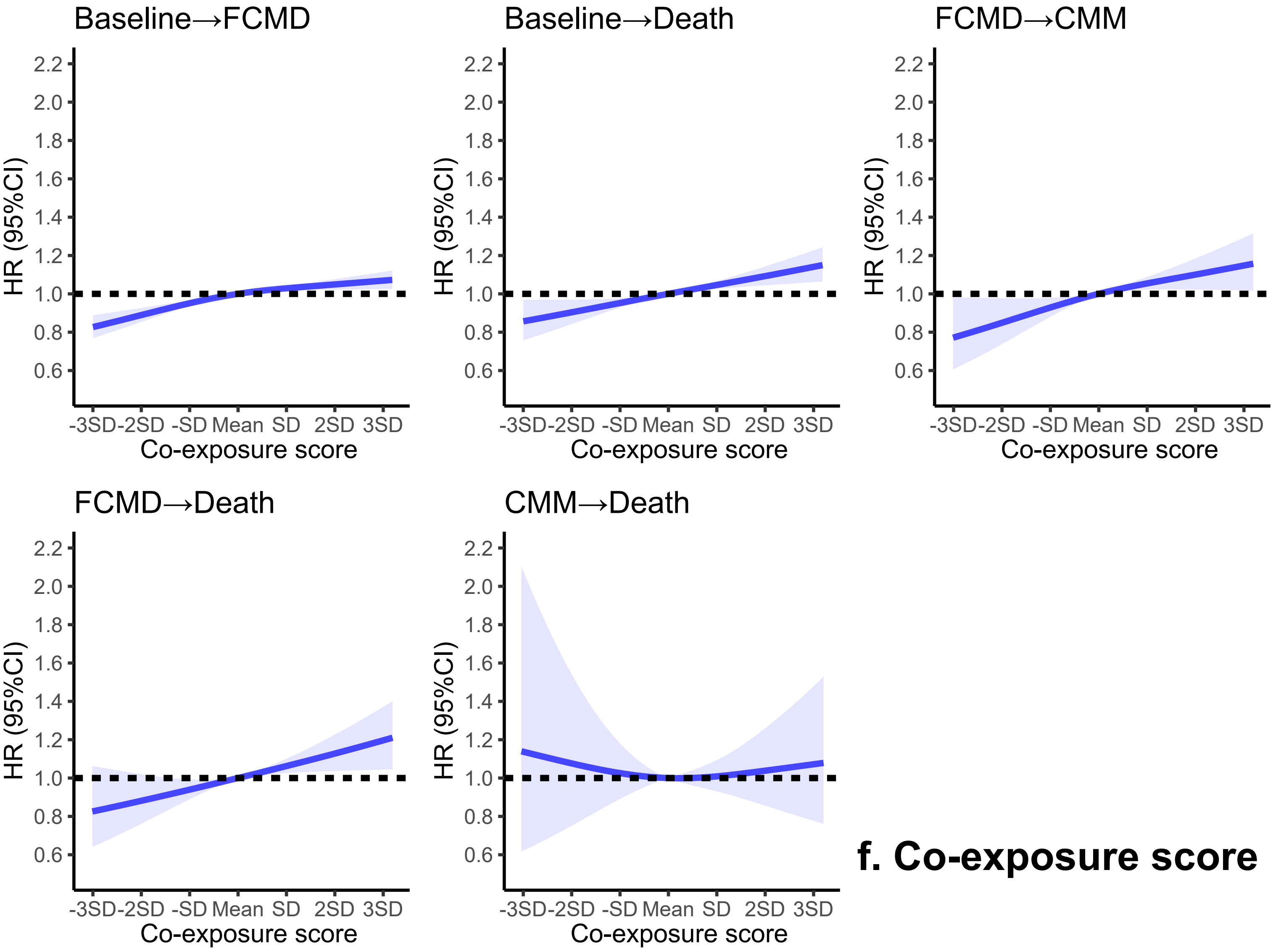


**f. Co-exposure score**

### Figure S4

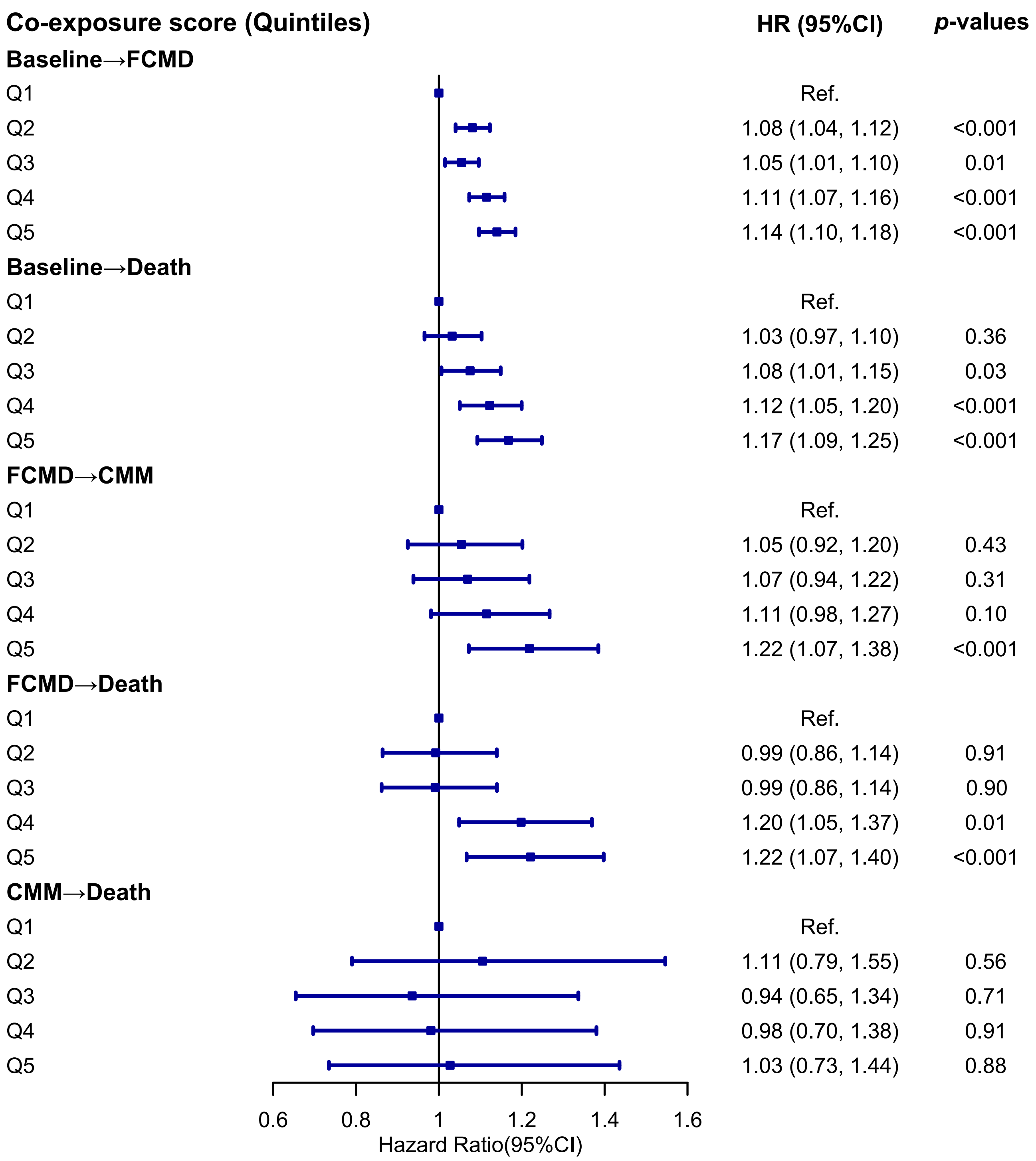
