## Supplement methods for "Long-term air pollution, cardiometabolic multimorbidity, and genetic susceptibility: a multi-state modeling study of 415,855 participants"

*Land Use Regression (LUR) model*

Air pollution estimates for the year 2010 were modelled for each address using a Land Use Regression (LUR) model developed as part of the European Study of Cohorts for Air Pollution Effects. Air pollution estimates for the years 2005-2007 were derived from EU-wide air pollution maps using another LUR. LUR models calculate the spatial variation of annual moving average air pollutant concentration using the predictor variables obtained from the Geographic Information System such as traffic, land use, and topography by a resolution of 100m * 100m. Participants’ ambient air pollution concentrations were then assigned according to their coordinates in the 100m * 100m grid cells. Previous reports with leave-one-out cross-validation showed good model performance for PM_2.5_, PM_10_, NO_2_ and NO_x_ (R^2^ of cross-validation =77%, 88%, 87% and 88%, respectively) and a comparatively moderate performance for PM_coarse_ (cross-validation R^2^=57%) in the southeast England area (London/Oxford). Details on the development and validation of the ESCAPE LUR models have been described elsewhere [1-3]. The LUR estimates of PM were valid for 400 km from Greater London but not beyond, and as such, patients living in northern England and Scotland were eliminated from the PM analyses.

*Assessment of covariates*

Height and weight were measured by trained nurses during the baseline assessment center visit, and BMI was calculated by dividing weight in kilograms by the square of height in meters and then was classified into: underweight or normal weight (<25), overweight (25 to <30), and obese (≥30). Moderate alcohol intake status was defined as: male: <28g/day; female: <14g/day. High-level physical activity status was defined as: ≥150 min/week moderate or ≥75 min/week vigorous or 150 min/week mixed (moderate + vigorous) activity. Physical activity was assessed using the Metabolic Equivalent Task minutes based on adopted items from the short International Physical Activity Questionnaire [4]. Observations with any missing values of the covariates were excluded from this study.

*Genotyping information*

Details of the genotyping, data imputation, and quality control in UK Biobank have been previously reported [5, 6]. Briefly, approximately 10% of participants were genotyped using the Applied BiosystemsTM UK BiLEVE Axiom^TM^ Array by Affymetrix (807,411 sites), with the remaining participants being genotyped using the Applied Biosystems^TM^ UK Biobank Axiom Array (825,927 sites), both of which were specifically designed for UK Biobank with 95% shared sites. Phasing and imputation of the remaining 5% unique single nucleotide polymorphisms (SNPs) were conducted using SHAPEIT3 and IMPUTE2 with both the merged UK10K and 1000 Genomes Phase 3 reference panel and the Haplotype Reference Consortium (HRC) reference panel (i.e., array imputation). SNPs after the array imputation were considered for building genetic risk score (GRS). We directly used the imputed genetic data without further quality control [5].
