## Supplementary material for "Long-term air pollution, cardiometabolic multimorbidity, and genetic susceptibility: a multi-state modeling study of 415,855 participants": Table S1-S6&S8-S15

**Table of contents**

**Table S1** Characteristics of genetic variants associated with type II diabetes, ischemic heart disease, and stroke

**Table S2** Baseline characteristics of 415,855 participants by incident disease status during follow-up

**Table S3** Associations of five air pollutants with the risks of first cardiometabolic disease, cardiometabolic multimorbidity, and mortality using Cox model

**Table S4** Associations of the co-exposure score based on PM_2.5_, NO_2_, and NO_x_ with the trajectories of cardiometabolic multimorbidity of pattern A using multi-state model

**Table S5** Associations of air pollution with the trajectories of cardiometabolic multimorbidity of pattern B using multi-state model

**Table S6** Associations of categorical co-exposure score (quintiles) with the trajectories of cardiometabolic multimorbidity of pattern B using multi-state model

**Table S7** Associations of CMD-related genetic variants with the trajectories of cardiometabolic multimorbidity of pattern A using multi-state model (a-c) and their interactions with co-exposure score (d-f)

**Table S8** Associations of weighted genetic risk score with the trajectories of cardiometabolic multimorbidity of pattern A using multi-state model

**Table S9** Mutual associations of co-exposure score and genetic risk score with the trajectories of cardiometabolic multimorbidity of pattern A using multi-state model in model with and without interaction terms

**Table S10** Joint associations of weighted genetic risk score and co-exposure score of five transition states on pattern A using multi-state model

**Table S11** Associations of air pollution with the trajectories of cardiometabolic multimorbidity of pattern A using multi-state model additionally adjusted for baseline diet behaviors, cholesterol levels, and blood pressure

**Table S12** Associations of air pollution with the trajectories of cardiometabolic multimorbidity of pattern A using multi-state model additionally adjusted for genetic principal components

**Table S13** Associations of air pollution with the trajectories of cardiometabolic multimorbidity of pattern A using multi-state model in white participants

**Table S14** Associations of air pollution with the trajectories of cardiometabolic multimorbidity of pattern A using multi-state model in participants living in the baseline address for more than five years

**Table S15** Associations of air pollution with the trajectories of cardiometabolic multimorbidity of pattern A using multi-state model by the age of having the first cardiometabolic disease

### **Table S1** Characteristics of genetic variants associated with type II diabetes, ischemic heart disease, and stroke ^1-3^

| **Trait** | **SNP** | **CHR** | **Genes** | ***p-*values** | **Effect allele** | **Non-effect allele** | **OR** | **Frequency** |
| --- | --- | --- | --- | --- | --- | --- | --- | --- |
| T2D | rs17106184 | 1 | *FAF1* | 4.00×10^-9^ | A | G | 1.1 | 0.92 |
|  | **rs10923931** | 1 | *NOTCH2* | 4.00×10^-8^ | T | G | 1.13 | 0.11 |
|  | **rs340874** | 1 | *PROX1* | 7.00×10^-10^ | C | T | 1.07 | 0.52 |
|  | rs780094 | 2 | *GCKR* | 1.00×10^-9^ | C | T | 1.06 | 0.39 |
|  | **rs7578597** | 2 | *THADA* | 1.00×10^-9^ | C | T | 1.15 | 0.90 |
|  | rs243021 | 2 | *BCL11A* | 3.00×10^-15^ | A | G | 1.08 | 0.50 |
|  | rs7560163 | 2 | *RBM43*, *RND3* | 7.00×10^-9^ | G | C | 1.33 | 0.86 |
|  | rs7593730 | 2 | *ITGB6*, *RBMS1* | 4.00×10^-8^ | C | T | 1.11 | 0.78 |
|  | **rs3923113** | 2 | *GRB14* | 1.00×10^-8^ | C | A | 1.09 | 0.74 |
|  | **rs7578326** | 2 | *IRS1* | 5.00×10^-20^ | G | A | 1.11 | 0.71 |
|  | **rs1801282** | 3 | *PPARG* | 6.00×10^-10^ | G | C | 1.16 | 0.88 |
|  | **rs7612463** | 3 | *UBE2E2* | 7.00×10^-9^ | A | C | 1.1 | 0.87 |
|  | **rs831571** | 3 | *PSMD6* | 8.00×10^-11^ | C | T | 1.09 | 0.61 |
|  | **rs4607103** | 3 | *ADAMTS9* | 1.00×10^-8^ | T | C | 1.09 | 0.76 |
|  | **rs11717195** | 3 | *ADCY5* | 2.00×10^-8^ | C | T | 1.09 | 0.78 |
|  | **rs4402960** | 3 | *IGF2BP2* | 2.00×10^-9^ | T | G | 1.17 | 0.29 |
|  | rs16861329 | 3 | *ST6GAL1* | 3.00×10^-8^ | T | C | 1.09 | 0.75 |
|  | **rs6808574** | 3 | *LPP* | 6.00×10^-9^ | C | T | 1.07 | 0.79 |
|  | **rs6815464** | 4 | *MAEA* | 2.00×10^-20^ | G | C | 1.13 | 0.58 |
|  | **rs1801214** | 4 | *WFS1* | 3.00×10^-8^ | T | C | 1.13 | 0.73 |
|  | rs6813195 | 4 | *TMEM154* | 4.00×10^-14^ | T | C | 1.08 | 0.59 |
|  | **rs702634** | 5 | *ARL15* | 7.00×10^-9^ | A | G | 1.06 | 0.76 |
|  | **rs4457053** | 5 | *ZBED3* | 3.00×10^-12^ | A | G | 1.08 | 0.20 |
|  | rs9502570 | 6 | *RREB1*, *SSR1* | 1.00×10^-9^ | T | C | 1.06 | 0.45 |
|  | **rs7754840** | 6 | *CDKAL1* | 4.00×10^-11^ | C | G | 1.12 | 0.36 |
|  | **rs3132524** | 6 | *TCF19*, *POU5F1* | 4.00×10^-9^ | C | T | 1.07 | 0.38 |
|  | rs9470794 | 6 | *ZFAND3* | 2.00×10^-10^ | C | T | 1.12 | 0.27 |
|  | rs1535500 | 6 | *KCNK16* | 2.00×10^-8^ | T | G | 1.08 | 0.42 |
|  | rs1048886 | 6 | *C6orf57* | 3.00×10^-8^ | G | A | 1.54 | 0.18 |
|  | rs2191349 | 7 | *DGKB*, *TMEM195* | 1.00×10^-8^ | C | T | 1.06 | 0.52 |
|  | **rs864745** | 7 | *JAZF1* | 5.00×10^-14^ | A | G | 1.1 | 0.50 |
|  | rs4607517 | 7 | *GCK* | 5.00×10^-8^ | G | A | 1.07 | 0.16 |
|  | rs6467136 | 7 | *PAX4*, *GCC1* | 5.00×10^-11^ | G | A | 1.11 | 0.79 |
|  | rs791595 | 7 | *MIR129*, *LEP* | 3.00×10^-13^ | G | A | 1.17 | 0.08 |
|  | rs972283 | 7 | *KLF14* | 2.00×10^-10^ | G | A | 1.07 | 0.69 |
|  | rs515071 | 8 | *ANK1* | 1.00×10^-8^ | C | T | 1.18 | 0.79 |
|  | rs896854 | 8 | *TP53INP1* | 1.00×10^-9^ | T | C | 1.06 | 0.24 |
|  | **rs13266634** | 8 | *SLC30A8* | 5.00×10^-8^ | G | A | 1.12 | 0.69 |
|  | **rs7041847** | 9 | *GLIS3* | 2.00×10^-14^ | T | C | 1.1 | 0.41 |
|  | rs17584499 | 9 | *PTPRD* | 9.00×10^-10^ | C | T | 1.57 | 0.06 |
|  | rs10811661 | 9 | *CDKN2A*, *CDKN2B* | 5.00×10^-8^ | T | C | 1.2 | 0.83 |
|  | **rs13292136** | 9 | *CHCHD9* | 3.00×10^-8^ | A | G | 1.11 | 0.90 |
|  | rs11787792 | 9 | *GPSM1* | 2.00×10^-10^ | G | A | 1.15 | 0.87 |
|  | **rs12779790** | 10 | *CDC123*, *CAMK1D* | 1.00×10^-10^ | T | C | 1.11 | 0.18 |
|  | **rs1802295** | 10 | *VPS26A* | 4.00×10^-8^ | G | A | 1.08 | 0.26 |
|  | rs12571751 | 10 | *ZMIZ1* | 2.00×10^-10^ | T | C | 1.09 | 0.51 |
|  | **rs1111875** | 10 | *HHEX* | 6.00×10^-10^ | T | C | 1.13 | 0.52 |
|  | **rs7903146** | 10 | *TCF7L2* | 2.00×10^-34^ | T | C | 1.65 | 0.30 |
|  | rs10886471 | 10 | *GRK5* | 7.00×10^-9^ | A | G | 1.12 | 0.78 |
|  | **rs3842770** | 11 | *INS-IGF2* | 3.00×10^-8^ | T | C | 1.14 | 0.23 |
|  | **rs2237892** | 11 | *KCNQ1* | 2.00×10^-42^ | T | C | 1.4 | 0.61 |
|  | **rs5215** | 11 | *KCNJ11* | 5.00×10^-11^ | C | A | 1.14 | 0.27 |
|  | rs1552224 | 11 | *CENTD2* | 1.00×10^-22^ | T | C | 1.14 | 0.45 |
|  | **rs1387153** | 11 | *MTNR1B* | 8.00×10^-15^ | C | G | 1.09 | 0.35 |
|  | **rs1531343** | 12 | *HMGA2* | 4.00×10^-9^ | T | C | 1.1 | 0.21 |
|  | **rs7961581** | 12 | *LGR5*, *TSPAN8* | 1.00×10^-9^ | A | T | 1.09 | 0.27 |
|  | **rs7957197** | 12 | *HNF1A* | 2.00×10^-8^ | G | C | 1.07 | 0.89 |
|  | **rs1727313** | 12 | *MPHOSPH9* | 1.00×10^-8^ | A | G | 1.06 | 0.50 |
|  | rs9552911 | 13 | *SGCG*, *SACS* | 2.00×10^-8^ | A | G | 1.49 | 0.93 |
|  | **rs1359790** | 13 | *SPRY2* | 6.00×10^-9^ | C | T | 1.15 | 0.71 |
|  | **rs7403531** | 15 | *RASGRP1* | 4.00×10^-9^ | G | T | 1.1 | 0.35 |
|  | rs7172432 | 15 | *C2CD4B*, *C2CD4A* | 9.00×10^-14^ | G | A | 1.11 | 0.58 |
|  | **rs7178572** | 15 | *HMG20A* | 1.00×10^-8^ | G | A | 1.11 | 0.70 |
|  | rs11634397 | 15 | *ZFAND6* | 2.00×10^-9^ | G | A | 1.06 | 0.44 |
|  | rs2028299 | 15 | *AP3S2* | 2.00×10^-11^ | A | C | 1.1 | 0.31 |
|  | rs8042680 | 15 | *PRC1* | 2.00×10^-10^ | A | C | 1.07 | 0.74 |
|  | rs8050136 | 16 | *FTO* | 1.00×10^-12^ | A | C | 1.17 | 0.38 |
|  | rs391300 | 17 | *SRR* | 3.00×10^-9^ | C | T | 1.28 | 0.62 |
|  | rs312457 | 17 | *SLC16A13* | 8.00×10^-13^ | A | G | 1.2 | 0.08 |
|  | **rs4430796** | 17 | *HNF1B* | 2.00×10^-11^ | A | G | 1.19 | 0.28 |
|  | rs8090011 | 18 | *LAMA1* | 8.0010×^-9^ | G | C | 1.13 | 0.38 |
|  | rs12970134 | 18 | *MC4R* | 3.00×10^-8^ | A | G | 1.08 | 0.27 |
|  | rs3786897 | 19 | *PEPD* | 1.00×10^-8^ | G | A | 1.1 | 0.56 |
|  | rs4812829 | 20 | *HNF4A* | 3.00×10^-10^ | A | G | 1.09 | 0.29 |
| IHD | **rs11206510** | 1 | *PCSK9* | 2.34×10^-8^ | T | C | 1.08 | 0.85 |
|  | **rs17114036** | 1 | *PPAP2B* | 5.00×10^14^ | A | G | 1.13 | 0.92 |
|  | rs646776 | 1 | *SORT1* | 1.97×10^-23^ | T | C | 1.11 | 0.75 |
|  | rs4845625 | 1 | *IL6R* | 2.60×10^-9^ | T | C | 1.05 | 0.45 |
|  | **rs17464857** | 1 | *MIA3****** | 1.01×10^-12^ | T | G | 1.06 | 0.86 |
|  | **rs17465637** | 1 | *MIA3****** | 1.01×10^-12^ | C | A | 1.08 | 0.66 |
|  | rs16986953 | 2 | *AK097927* | 1.45×10^-8^ | A | G | 1.09 | 0.10 |
|  | rs515135 | 2 | *APOB* | 2.89×10^-8^ | C | T | 1.07 | 0.79 |
|  | **rs6544713** | 2 | *ABCG8* | 2.60×10^-8^ | T | C | 1.05 | 0.32 |
|  | **rs1561198** | 2 | *VAMP5* | 3.62×10^-10^ | T | C | 1.06 | 0.46 |
|  | **rs2252641** | 2 | *TEX41* | 3.00×10^-9^ | C | T | 1.03 | 0.48 |
|  | **rs6725887** | 2 | *WDR12* | 2.15×10^-18^ | C | T | 1.14 | 0.11 |
|  | rs9818870 | 3 | *MRAS* | 2.89×10^-9^ | T | C | 1.07 | 0.14 |
|  | **rs1878406** | 4 | *EDNRA****** | 8.82×10^-10^ | T | C | 1.06 | 0.16 |
|  | **rs7692387** | 4 | *GUCY1A3* | 6.07×10^-9^ | G | A | 1.07 | 0.81 |
|  | rs273909 | 5 | *SLC22A4* | 1.24×10^-4^ | G | A | 1.06 | 0.12 |
|  | rs6903956 | 6 | *ADTRP* | 0.96 | A | G | 1.00 | 0.35 |
|  | rs12526453 | 6 | *PHACTR1* | 1.81×10^-42^ | C | G | 1.10 | 0.71 |
|  | rs17609940 | 6 | *ANKS1A* | 0.03 | G | C | 1.03 | 0.82 |
|  | **rs10947789** | 6 | *KCNK5* | 1.85×10^-8^ | T | C | 1.05 | 0.78 |
|  | **rs12190287** | 6 | *TCF21* | 1.98×10^-11^ | C | G | 1.06 | 0.62 |
|  | **rs2048327** | 6 | *SLC22A3* | 5.39×10^-39^ | C | T | 1.06 | 0.35 |
|  | **rs3798220** | 6 | *LPA* | 5.39×10^-39^ | C | T | 1.42 | 0.02 |
|  | rs4252120 | 6 | *PLG* | 1.64×10^-32^ | T | C | 1.03 | 0.74 |
|  | rs2023938 | 7 | *HDAC9* | 8.05×10^-11^ | C | T | 1.06 | 0.10 |
|  | rs10953541 | 7 | *7q22* | 1.02×10^-5^ | C | T | 1.05 | 0.78 |
|  | rs11556924 | 7 | *ZC3HC1* | 5.34×10^-11^ | C | T | 1.08 | 0.69 |
|  | **rs264** | 8 | *LPL* | 1.06×10^-5^ | G | A | 1.06 | 0.85 |
|  | **rs2954029** | 8 | *TRIB1* | 2.61×10^-6^ | A | T | 1.04 | 0.55 |
|  | **rs3217992** | 9 | *CDKN2B-AS1****** | 2.29×10^-98^ | T | C | 1.14 | 0.39 |
|  | **rs4977574** | 9 | *CDKN2B-AS1****** | 2.29×10^-98^ | G | A | 1.21 | 0.49 |
|  | **rs579459** | 9 | *ABO****** | 1.19×10^-11^ | C | T | 1.08 | 0.21 |
|  | rs2505083 | 10 | *KIAA1462* | 4.41×10^-11^ | C | T | 1.06 | 0.40 |
|  | **rs2047009** | 10 | *CXCL12* | 5.55×10^-15^ | G | T | 1.06 | 0.48 |
|  | rs501120 | 10 | *CXCL12* | 5.55×10^-15^ | T | C | 1.08 | 0.81 |
|  | rs11203042 | 10 | *LIPA* | 5.15×10^-12^ | T | C | 1.04 | 0.45 |
|  | **rs1412444** | 10 | *LIPA* | 5.15×10^-12^ | T | C | 1.07 | 0.37 |
|  | **rs12413409** | 10 | *CNNM2* | 4.65×10^-9^ | G | A | 1.08 | 0.89 |
|  | rs974819 | 11 | *PDGFD* | 7.05×10^-11^ | T | C | 1.07 | 0.33 |
|  | **rs964184** | 11 | *ZPR1* | 5.60×10^-5^ | G | C | 1.05 | 0.18 |
|  | rs7136259 | 12 | *ATP2B1* | 6.17×10^-11^ | T | C | 1.04 | 0.43 |
|  | **rs3184504*** | 12 | *SH2B3* | 1.03×10^-9^ | T | C | 1.07 | 0.42 |
|  | rs9319428 | 13 | *FLT1* | 7.13×10^-5^ | A | G | 1.04 | 0.31 |
|  | rs4773144 | 13 | *COL4A1/A2* | 1.83×10^-10^ | G | A | 1.05 | 0.43 |
|  | **rs9515203** | 13 | *COL4A1/A2* | 1.83×10^-10^ | T | C | 1.07 | 0.76 |
|  | **rs2895811** | 14 | *HHIPL1* | 1.38×10^-8^ | C | T | 1.04 | 0.41 |
|  | **rs7173743** | 15 | *ADAMTS7* | 4.44×10^-16^ | T | C | 1.08 | 0.56 |
|  | **rs17514846** | 15 | *FURIN****** | 3.10×10^-7^ | A | C | 1.05 | 0.44 |
|  | rs216172 | 17 | *SMG6* | 5.07×10^-7^ | C | G | 1.05 | 0.35 |
|  | rs12936587 | 17 | *RAI1-PEMT-RASD1* | 8.24×10^-4^ | G | A | 1.03 | 0.61 |
|  | **rs46522** | 17 | *UBE2Z* | 1.84×10^-5^ | T | C | 1.04 | 0.51 |
|  | rs1122608 | 19 | *LDLR* | 4.44×10^-15^ | G | T | 1.08 | 0.77 |
|  | **rs2075650** | 19 | *TOMM40* | 7.07×10^-11^ | G | A | 1.07 | 0.13 |
|  | **rs445925** | 19 | *APOC1* | 7.07×10^-11^ | G | A | 1.09 | 0.90 |
|  | **rs9982601** | 21 | *KCNE2 (gene desert)* | 1.33×10^-15^ | T | C | 1.12 | 0.13 |
| Stroke | rs880315 | 1 | *CASZ1* | 3.62 ×10^−10^ | C | T | 1.05 | 0.40 |
|  | rs12037987 | 1 | *WNT2B* | 2.73 ×10^−8^ | C | T | 1.07 | 0.16 |
|  | rs146390073 | 1 | *RGS7* | 2.20×10^−8^ | T | C | 1.95 | 0.02 |
|  | rs12476527 | 2 | *KCNK3* | 6.44×10^−8^ | G | T | 1.05 | 0.48 |
|  | **rs7610618** | 3 | *TM4SF4–TM4Sn* | 1.44×10^−8^ | T | C | 2.33 | 0.01 |
|  | rs34311906 | 4 | *ANK2* | 1.07×10^−8^ | C | T | 1.07 | 0.41 |
|  | **rs17612742** | 4 | *EDNRA****** | 1.46 ×10^−11^ | C | T | 1.19 | 0.21 |
|  | **rs6825454** | 4 | *FGA* | 7.43 ×10^−10^ | C | T | 1.06 | 0.31 |
|  | rs11957829 | 5 | *LOC100505841* | 7.51 ×10^−9^ | A | G | 1.07 | 0.82 |
|  | rs6891174 | 5 | *NKX2-5* | 5.82 ×10^−9^ | A | G | 1.11 | 0.35 |
|  | rs16896398 | 6 | *SLC22A7–ZNF318* | 1.30×10^−8^ | T | A | 1.05 | 0.34 |
|  | **rs42039** | 7 | *CDK6* | 6.55×10^−9^ | C | T | 1.07 | 0.77 |
|  | **rs7859727** | 9 | *CDKN2B-AS1****** | 4.22 ×10^−10^ | T | C | 1.05 | 0.53 |
|  | rs10820405 | 9 | *LINC01492* | 4.51 ×10^−8^ | G | A | 1.2 | 0.82 |
|  | **rs2295786** | 10 | *SH3PXD2A* | 1.80×10^−10^ | A | T | 1.05 | 0.60 |
|  | **rs7304841** | 12 | *PDE3A* | 4.93 ×10^−8^ | A | C | 1.05 | 0.59 |
|  | rs35436 | 12 | *TBX3* | 2.87×10^−8^ | C | T | 1.05 | 0.62 |
|  | **rs9526212** | 13 | *LRCH1* | 5.03×10^−10^ | G | A | 1.06 | 0.76 |
|  | **rs4932370** | 15 | *FURIN****** | 2.88×10^−8^ | A | G | 1.05 | 0.33 |
|  | rs11867415 | 17 | *PRPF8* | 4.81 ×10^−8^ | G | A | 1.09 | 0.18 |
|  | **rs2229383** | 19 | *ILF3* | 4.72 ×10^−8^ | T | G | 1.05 | 0.65 |
|  | rs8103309 | 19 | *SMARCA4-LDLR* | 3.40×10^−8^ | T | C | 1.05 | 0.65 |
|  | **rs12124533** | 1 | *TSPAN2* | 1.22 ×10^−8^ | T | C | 1.17 | 0.24 |
|  | rs1052053 | 1 | *PMF1*–*SEMA4A* | 2.70×10^−14^ | G | A | 1.06 | 0.40 |
|  | **rs13143308** | 4 | *PITX2* | 1.86×10^−47^ | T | G | 1.32 | 0.28 |
|  | rs4959130 | 6 | *FOXF2* | 1.42 ×10^−9^ | A | G | 1.08 | 0.14 |
|  | **rs2107595** | 7 | *HDAC9–TWIST1* | 3.65×10^−15^ | A | G | 1.21 | 0.24 |
|  | **rs635634** | 9 | *ABO****** | 9.18×10^−9^ | T | C | 1.08 | 0.19 |
|  | rs2005108 | 11 | *MMP12* | 3.33×10^−8^ | T | C | 1.08 | 0.12 |
|  | **rs3184504*** | 12 | *SH2B3****** | 2.17×10^−14^ | T | C | 1.08 | 0.45 |
|  | rs12932445 | 16 | *ZFHX3* | 6.86×10^−18^ | C | T | 1.2 | 0.21 |
|  | rs12445022 | 16 | *ZCCHC14* | 1.05×10^−10^ | A | G | 1.06 | 0.31 |

Bolded: selected to build genetic risk score (detail see Table S6)

***:** overlapped SNPs or genes (for selected SNPs only)

**Table S2** Baseline characteristics of 415,855 participants by incident disease status during follow-up

| **Characteristic^a^** | **Total**  **N =415855** | **Participants with FCMD**  **N=33375** | **Participants with CMM**  **N=3257** |
| --- | --- | --- | --- |
| **Sex (male), %** | 231115 (55.58 %) | 13329 (39.94 %) | 1054 (32.36 %) |
| **BMI (kg/m^2^)** | 27.25 (4.69) | 29.59 (± 5.42) | 31.03 (± 5.56) |
| **Age at baseline, year** | 56.23 (8.09) | 60.10 (± 6.94) | 61.09 (± 6.53) |
| **Smoking status, %** |  |  |  |
| Never smoker | 231696 (55.72 %) | 15098 (45.24 %) | 1290 (39.61 %) |
| Former smoker | 142138 (34.18 %) | 13740 (41.17 %) | 1471 (45.16 %) |
| Current smoker | 42021 (10.10 %) | 4537 (13.59 %) | 496 (15.23 %) |
| **Years of education (≥ 10 years), %** | 276369 (66.46 %) | 17964 (53.82 %) | 1574 (48.33 %) |
| **Moderate alcohol intake^b^ (yes), %** | 207782 (49.97 %) | 15332 (45.94 %) | 1390 (42.68 %) |
| **High-level physical activity^c^ (yes), %** | 298231 (71.72 %) | 21900 (65.62 %) | 1997 (61.31 %) |
| **Total household income (≥** ₤**31,000), %** | 167654 (40.32 %) | 17396 (52.12 %) | 1830 (56.19 %) |
| **Employment status (employed), %** | 246624 (59.31 %) | 14253 (42.71 %) | 1239 (38.04 %) |
| **White, %** | 394348 (94.83 %) | 31090 (93.15 %) | 2920 (89.65 %) |

CMDs: cardiometabolic diseases; FCMD: first cardiometabolic disease; CMM: cardiometabolic multimorbidity.

a: Results are presented as mean (standard deviation) for continuous variables or number (percentage) for categorical variables;

b: Moderate alcohol intake: male: <28g/day; female: <14g/day;

c: High-level physical activity: ≥150 min/week moderate or ≥75 min/week vigorous or 150 min/week mixed (moderate + vigorous) activity;

### Table S3 Associations of five air pollutants with the risks of first cardiometabolic disease, cardiometabolic multimorbidity, and mortality using Cox model

| **Air pollutants** | **First cardiometabolic disease** | | **Cardiometabolic multimorbidity** | | **Mortality** | |
| --- | --- | --- | --- | --- | --- | --- |
|  | **HR (95% CI)** | ***p-*values** | **HR (95% CI)** | ***p-*values** | **HR (95% CI)** | ***p-*values** |
| **PM_2.5_** | 1.26 (1.20 - 1.34) | <0.0001 | 1.68 (1.41 - 1.99) | <0.0001 | 1.02 (1.02 - 1.03) | <0.0001 |
| **PM_2.5-10_** | 1.04 (0.97 - 1.11) | 0.29 | 1.10 (0.90 - 1.36) | 0.35 | 1.01 (0.92 - 1.12) | 0.82 |
| **PM_10_** | 1.03 (1.00 - 1.06) | 0.09 | 1.14 (1.03 - 1.26) | 0.01 | 1.14 (1.08 - 1.19) | <0.0001 |
| **NO_2_** | 1.02 (1.01 - 1.02) | <0.0001 | 1.05 (1.03 - 1.07) | <0.0001 | 1.04 (1.03 - 1.05) | <0.0001 |
| **NO_x_** | 1.01 (1.01 - 1.02) | <0.0001 | 1.03 (1.02 - 1.04) | <0.0001 | 1.02 (1.02 - 1.03) | <0.0001 |

a: Models adjusted for age, sex, BMI, years of education, smoking status, moderate alcohol intake, high-level physical activity, total household income, and employment status. Estimates of air pollutants were demonstrated per 5-μg/m^3^ increase.

### Table S4 Associations of the co-exposure score based on PM_2.5_, NO_2_, and NO_x_ with the trajectories of cardiometabolic multimorbidity of pattern A using multi-state model ^a^

| **Trajectories** | **HR (95%CI)** | ***p-*values** |
| --- | --- | --- |
| **Baseline→FCMD** | 1.05 (1.03, 1.06) | <0.0001 |
| **Baseline→Death** | 1.06 (1.04, 1.08) | <0.0001 |
| **FCMD→CMM** | 1.07 (1.03, 1.11) | <0.0001 |
| **FCMD→Death** | 1.08 (1.04, 1.12) | <0.0001 |
| **CMM→Death** | 1.00 (0.91, 1.11) | 0.97 |

a: Models adjusted for age, sex, ethnicity, BMI, years of education, smoking status, moderate alcohol intake, high-level physical activity, total household income, and employment status. Estimates were demonstrated per one SD increase in the co-exposure score.

### Table S5 Associations of air pollution with the trajectories of cardiometabolic multimorbidity of pattern B using multi-state model ^a^

| **Air pollutants** | **HR (95%CI)** | ***p-*values** | **Air pollutants** | **HR (95%CI)** | ***p-*values** |
| --- | --- | --- | --- | --- | --- |
| **PM_2.5_** |  |  | **NO_2_** |  |  |
| Baseline→IHD | 1.13 (1.04, 1.22) | <0.0001 | Baseline→IHD | 0.98 (0.93, 1.02) | 0.34 |
| Baseline→T2D | 1.47 (1.34, 1.60) | <0.0001 | Baseline→T2D | 1.21 (1.15, 1.28) | <0.0001 |
| Baseline→Stroke | 1.23 (1.02, 1.48) | 0.03 | Baseline→Stroke | 1.04 (0.93, 1.16) | 0.47 |
| Baseline→ Death | 1.29 (1.17, 1.42) | <0.0001 | Baseline→ Death | 1.15 (1.09, 1.22) | <0.0001 |
| IHD→CMM | 1.51 (1.08, 2.10) | 0.02 | IHD→CMM | 1.28 (1.04, 1.58) | 0.02 |
| IHD→Death | 1.10 (1.04, 1.17) | <0.0001 | IHD→Death | 1.01 (1.01, 1.02) | <0.0001 |
| T2D→CMM | 1.05 (1.01, 1.10) | 0.02 | T2D→CMM | 1.00 (1.00, 1.01) | 0.17 |
| T2D→Death | 1.03 (0.97, 1.10) | 0.36 | T2D→Death | 1.00 (1.00, 1.01) | 0.21 |
| Stroke→CMM | 1.01 (0.88, 1.15) | 0.94 | Stroke→CMM | 1.00 (0.98, 1.01) | 0.72 |
| Stroke→Death | 1.01 (0.92, 1.10) | 0.84 | Stroke→Death | 1.01 (1.00, 1.02) | 0.16 |
| CMM→Death | 0.97 (0.88, 1.06) | 0.50 | CMM→Death | 1.00 (0.99, 1.01) | 0.63 |
| **PM_2.5-10_** |  |  | **NO_x_** |  |  |
| Baseline→IHD | 0.76 (0.48, 1.21) | 0.24 | Baseline→IHD | 1.03 (1.00, 1.05) | 0.05 |
| Baseline→T2D | 1.86 (1.12, 3.08) | 0.02 | Baseline→T2D | 1.12 (1.09, 1.15) | <0.0001 |
| Baseline→Stroke | 1.89 (0.65, 5.51) | 0.24 | Baseline→Stroke | 1.05 (0.98, 1.11) | 0.14 |
| Baseline→ Death | 1.18 (0.67, 2.08) | 0.57 | Baseline→ Death | 1.09 (1.06, 1.13) | <0.0001 |
| IHD→CMM | 0.90 (0.12, 6.56) | 0.92 | IHD→CMM | 1.15 (1.04, 1.27) | 0.01 |
| IHD→Death | 0.95 (0.88, 1.02) | 0.16 | IHD→Death | 1.01 (1.00, 1.01) | <0.0001 |
| T2D→CMM | 1.01 (0.96, 1.06) | 0.73 | T2D→CMM | 1.00 (1.00, 1.00) | 0.15 |
| T2D→Death | 0.99 (0.92, 1.07) | 0.78 | T2D→Death | 1.00 (1.00, 1.01) | 0.34 |
| Stroke→CMM | 0.92 (0.78, 1.09) | 0.33 | Stroke→CMM | 1.00 (0.99, 1.01) | 0.78 |
| Stroke→Death | 0.94 (0.84, 1.04) | 0.23 | Stroke→Death | 1.00 (0.99, 1.01) | 0.98 |
| CMM→Death | 1.02 (0.92, 1.14) | 0.73 | CMM→Death | 1.00 (1.00, 1.01) | 0.54 |
| **PM_10_** |  |  | **Co-exposure score** |  |  |
| Baseline→IHD | 0.76 (0.61, 0.95) | 0.01 | Baseline→IHD | 1.01 (1.00, 1.03) | 0.13 |
| Baseline→T2D | 1.79 (1.40, 2.30) | <0.0001 | Baseline→T2D | 1.08 (1.06, 1.10) | <0.0001 |
| Baseline→Stroke | 1.26 (0.75, 2.11) | 0.38 | Baseline→Stroke | 1.03 (0.99, 1.08) | 0.09 |
| Baseline→ Death | 1.74 (1.32, 2.30) | <0.0001 | Baseline→ Death | 1.06 (1.04, 1.08) | <0.0001 |
| IHD→CMM | 1.24 (0.46, 3.35) | 0.67 | IHD→CMM | 1.10 (1.02, 1.18) | 0.01 |
| IHD→Death | 1.03 (1.00, 1.07) | 0.05 | IHD→Death | 1.13 (1.06, 1.20) | <0.0001 |
| T2D→CMM | 1.01 (0.99, 1.04) | 0.36 | T2D→CMM | 1.05 (1.00, 1.10) | 0.06 |
| T2D→Death | 1.01 (0.98, 1.05) | 0.53 | T2D→Death | 1.04 (0.97, 1.11) | 0.29 |
| Stroke→CMM | 0.98 (0.91, 1.06) | 0.58 | Stroke→CMM | 0.99 (0.85, 1.14) | 0.85 |
| Stroke→Death | 1.01 (0.96, 1.07) | 0.59 | Stroke→Death | 1.02 (0.93, 1.12) | 0.66 |
| CMM→Death | 1.03 (0.97, 1.09) | 0.32 | CMM→Death | 1.01 (0.91, 1.11) | 0.91 |

a: Models adjusted for age, sex, ethnicity, BMI, years of education, smoking status, moderate alcohol intake, high-level physical activity, total household income, and employment status. Estimates of air pollutants were demonstrated per 5-μg/m^3^ increase and estimates of co-exposure score were demonstrated per one SD increase.

### Table S6 Associations of categorical co-exposure score (quintiles) with the trajectories of cardiometabolic multimorbidity of pattern B using multi-state model ^a^

| **Co-exposure score** | **HR (95%CI)** | ***p-*values** |
| --- | --- | --- |
| **Baseline→IHD** |  |  |
| Q1 | Ref. | Ref. |
| Q2 | 1.06 (1.00, 1.11) | 0.04 |
| Q3 | 1.03 (0.97, 1.08) | 0.34 |
| Q4 | 1.08 (1.02, 1.13) | 0.01 |
| Q5 | 1.04 (0.99, 1.10) | 0.13 |
| **Baseline→ T2D** |  |  |
| Q1 | Ref. | Ref. |
| Q2 | 1.12 (1.05, 1.20) | <0.0001 |
| Q3 | 1.13 (1.06, 1.20) | <0.0001 |
| Q4 | 1.19 (1.12, 1.26) | <0.0001 |
| Q5 | 1.27 (1.19, 1.35) | <0.0001 |
| **Baseline→ Stroke** |  |  |
| Q1 | Ref. | Ref. |
| Q2 | 1.11 (0.98, 1.25) | 0.10 |
| Q3 | 0.96 (0.84, 1.09) | 0.53 |
| Q4 | 1.08 (0.95, 1.22) | 0.24 |
| Q5 | 1.18 (1.04, 1.34) | 0.01 |
| **Baseline→ Death** |  |  |
| Q1 | Ref. | Ref. |
| Q2 | 1.03 (0.97, 1.10) | 0.36 |
| Q3 | 1.08 (1.01, 1.15) | 0.03 |
| Q4 | 1.12 (1.05, 1.20) | <0.0001 |
| Q5 | 1.17 (1.09, 1.25) | <0.0001 |
| **IHD→CMM** |  |  |
| Q1 | Ref. | Ref. |
| Q2 | 0.88 (0.7, 1.11) | 0.27 |
| Q3 | 0.81 (0.64, 1.02) | 0.08 |
| Q4 | 0.89 (0.71, 1.12) | 0.32 |
| Q5 | 1.10 (0.87, 1.38) | 0.42 |
| **IHD→Death** |  |  |
| Q1 | Ref. | Ref. |
| Q2 | 1.01 (0.82, 1.25) | 0.89 |
| Q3 | 0.98 (0.79, 1.21) | 0.83 |
| Q4 | 1.39 (1.14, 1.70) | <0.0001 |
| Q5 | 1.38 (1.13, 1.70) | <0.0001 |
| **T2D→CMM** |  |  |
| Q1 | Ref. | Ref. |
| Q2 | 1.06 (0.90, 1.26) | 0.48 |
| Q3 | 1.17 (0.99, 1.38) | 0.07 |
| Q4 | 1.14 (0.97, 1.34) | 0.11 |
| Q5 | 1.19 (1.01, 1.40) | 0.04 |
| **T2D→Death** |  |  |
| Q1 | Ref. | Ref. |
| Q2 | 0.94 (0.75, 1.20) | 0.63 |
| Q3 | 1.02 (0.80, 1.28) | 0.89 |
| Q4 | 1.07 (0.85, 1.34) | 0.55 |
| Q5 | 1.09 (0.87, 1.37) | 0.46 |
| **Stroke→CMM** |  |  |
| Q1 | Ref. | Ref. |
| Q2 | 1.63 (1.01, 2.63) | 0.04 |
| Q3 | 1.22 (0.72, 2.07) | 0.45 |
| Q4 | 1.35 (0.82, 2.22) | 0.24 |
| Q5 | 1.34 (0.81, 2.21) | 0.26 |
| **Stroke→Death** |  |  |
| Q1 | Ref. | Ref. |
| Q2 | 0.96 (0.71, 1.28) | 0.76 |
| Q3 | 0.96 (0.71, 1.29) | 0.77 |
| Q4 | 0.96 (0.71, 1.29) | 0.76 |
| Q5 | 1.05 (0.78, 1.41) | 0.73 |
| **CMM→Death** |  |  |
| Q1 | Ref. | Ref. |
| Q2 | 1.11 (0.79, 1.55) | 0.56 |
| Q3 | 0.94 (0.65, 1.34) | 0.71 |
| Q4 | 0.98 (0.70, 1.38) | 0.91 |
| Q5 | 1.03 (0.73, 1.44) | 0.88 |

a: Models were adjusted for age, sex, ethnicity, BMI, years of education, smoking status, moderate alcohol intake, high-level physical activity, total household income, and employment status.

**Table S7** Associations of CMD-related genetic variants with the trajectories of cardiometabolic multimorbidity of pattern A using multi-state model (a-c) and the interactions of selected variants with co-exposure score (d-f)

(See attached .xlsx file)

### Table S8 Associations of weighted genetic risk score with the trajectories of cardiometabolic multimorbidity of pattern A using multi-state model ^a^

| **Trajectories** | **HR (95%CI)** | ***p-*values** |
| --- | --- | --- |
| **Baseline→FCMD** | 1.14 (1.13, 1.16) | <0.0001 |
| **Baseline→Death** | 0.98 (0.96, 1.00) | 0.04 |
| **FCMD→CMM** | 1.11 (1.07, 1.16) | <0.0001 |
| **FCMD→Death** | 0.96 (0.92, 1.00) | 0.04 |
| **CMM→Death** | 0.92 (0.83, 1.02) | 0.10 |

a: Models were adjusted for age, sex, ethnicity, BMI, years of education, smoking status, moderate alcohol intake, high-level physical activity, total household income, and employment status. Estimates were demonstrated per one SD increase in the weighted genetic risk score.

### Table S9 Mutual associations of co-exposure score and genetic risk score with the trajectories of cardiometabolic multimorbidity of pattern A using multi-state model in model with and without interaction terms

|  | **Trajectories** | **HR (95%CI)** | | | | ***p*-values of interaction** |
| --- | --- | --- | --- | --- | --- | --- |
|  |  | **co-exposure score** | ***p-*values** | **weighted genetic risk score** | ***p-*values** |  |
| **With interaction** | **Baseline→FCMD** | 1.05 (0.91, 1.21) | 0.52 | 1.15 (1.06, 1.24) | <0.0001 | 0.94 |
|  | **Baseline→Death** | 0.93 (0.72, 1.19) | 0.55 | 0.91 (0.79, 1.04) | 0.17 | 0.29 |
|  | **FCMD→CMM** | 1.45 (0.92, 2.27) | 0.11 | 1.31 (1.03, 1.67) | 0.03 | 0.19 |
|  | **FCMD→Death** | 0.95 (0.58, 1.55) | 0.83 | 0.89 (0.68, 1.17) | 0.41 | 0.62 |
|  | **CMM→Death** | 1.05 (0.91, 1.21) | 0.52 | 1.04 (0.55, 1.94) | 0.91 | 0.70 |
| **Without interaction** | **Baseline→FCMD** | 1.04 (1.03, 1.06) | <0.0001 | 1.14 (1.13, 1.16) | <0.0001 | - |
|  | **Baseline→Death** | 1.06 (1.04, 1.08) | <0.0001 | 0.98 (0.96, 1.00) | 0.04 | - |
|  | **FCMD→CMM** | 1.07 (1.03, 1.11) | <0.0001 | 1.11 (1.07, 1.16) | <0.0001 | - |
|  | **FCMD→Death** | 1.07 (1.03, 1.12) | <0.0001 | 0.96 (0.92, 1.00) | 0.03 | - |
|  | **CMM→Death** | 1.00 (0.90, 1.11) | 1.00 | 0.92 (0.83, 1.02) | 0.10 | - |

a: Models were adjusted for age, sex, ethnicity, BMI, years of education, smoking status, moderate alcohol intake, high-level physical activity, total household income, and employment status. Estimates were demonstrated per one SD increase in the co-exposure score and weighted genetic risk score.

### Table S10 Joint associations of weighted genetic risk score and co-exposure score of five transition states on pattern A using multi-state model ^a^

| **Trajectories** | **HR (95%CI)** | ***p-*values** |
| --- | --- | --- |
| **Baseline→FCMD** |  |  |
| Low and Q1 | Ref. | Ref. |
| Low and Q2 | 1.10 (1.04, 1.17) | <0.0001 |
| Low and Q3 | 1.03 (0.98, 1.10) | 0.25 |
| Low and Q4 | 1.11 (1.05, 1.18) | <0.0001 |
| Low and Q5 | 1.13 (1.07, 1.20) | <0.0001 |
| High and Q1 | 1.25 (1.18, 1.32) | <0.0001 |
| High and Q2 | 1.32 (1.25, 1.39) | <0.0001 |
| High and Q3 | 1.34 (1.27, 1.42) | <0.0001 |
| High and Q4 | 1.39 (1.32, 1.47) | <0.0001 |
| High and Q5 | 1.41 (1.34, 1.49) | <0.0001 |
| **Baseline→Death** |  |  |
| Low and Q1 | Ref. | Ref. |
| Low and Q2 | 1.00 (0.91, 1.10) | 0.99 |
| Low and Q3 | 1.07 (0.97, 1.17) | 0.16 |
| Low and Q4 | 1.04 (0.95, 1.15) | 0.37 |
| Low and Q5 | 1.14 (1.04, 1.25) | 0.01 |
| High and Q1 | 0.92 (0.84, 1.01) | 0.10 |
| High and Q2 | 0.99 (0.90, 1.09) | 0.85 |
| High and Q3 | 1.00 (0.91, 1.10) | 0.92 |
| High and Q4 | 1.11 (1.01, 1.22) | 0.03 |
| High and Q5 | 1.11 (1.01, 1.22) | 0.03 |
| **FCMD→CMM** |  |  |
| Low and Q1 | Ref. | Ref. |
| Low and Q2 | 1.05 (0.85, 1.29) | 0.66 |
| Low and Q3 | 1.17 (0.95, 1.43) | 0.14 |
| Low and Q4 | 1.15 (0.94, 1.40) | 0.18 |
| Low and Q5 | 1.40 (1.15, 1.70) | <0.0001 |
| High and Q1 | 1.27 (1.04, 1.55) | 0.02 |
| High and Q2 | 1.35 (1.12, 1.64) | <0.0001 |
| High and Q3 | 1.28 (1.05, 1.55) | 0.01 |
| High and Q4 | 1.38 (1.14, 1.66) | <0.0001 |
| High and Q5 | 1.40 (1.16, 1.70) | <0.0001 |
| **FCMD→Death** |  |  |
| Low and Q1 | Ref. | Ref. |
| Low and Q2 | 1.08 (0.88, 1.32) | 0.45 |
| Low and Q3 | 1.13 (0.92, 1.39) | 0.23 |
| Low and Q4 | 1.20 (0.98, 1.46) | 0.07 |
| Low and Q5 | 1.27 (1.04, 1.55) | 0.02 |
| High and Q1 | 1.01 (0.83, 1.24) | 0.90 |
| High and Q2 | 0.95 (0.77, 1.16) | 0.61 |
| High and Q3 | 0.92 (0.75, 1.13) | 0.41 |
| High and Q4 | 1.21 (1.00, 1.46) | 0.05 |
| High and Q5 | 1.20 (0.99, 1.46) | 0.06 |
| **CMM→Death** |  |  |
| Low and Q1 | Ref. | Ref. |
| Low and Q2 | 1.28 (0.76, 2.15) | 0.36 |
| Low and Q3 | 1.06 (0.62, 1.82) | 0.83 |
| Low and Q4 | 1.09 (0.65, 1.85) | 0.74 |
| Low and Q5 | 1.15 (0.69, 1.90) | 0.59 |
| High and Q1 | 0.99 (0.58, 1.69) | 0.97 |
| High and Q2 | 1.04 (0.63, 1.72) | 0.88 |
| High and Q3 | 0.89 (0.52, 1.51) | 0.66 |
| High and Q4 | 0.96 (0.58, 1.60) | 0.87 |
| High and Q5 | 0.94 (0.56, 1.56) | 0.81 |

a: Models were adjusted for age, sex, ethnicity, BMI, years of education, smoking status, moderate alcohol intake, high-level physical activity, total household income, and employment status.

### Table S11 Associations of air pollution with the trajectories of cardiometabolic multimorbidity of pattern A using multi-state model additionally adjusted for baseline diet behaviors, cholesterol levels, and blood pressure ^a^

|  | **HR (95%CI)** | ***p-*values** |
| --- | --- | --- |
| **PM_2.5_** |  |  |
| **Baseline→FCMD** | 1.22 (1.14, 1.29) | <0.0001 |
| **Baseline→Death** | 1.23 (1.10, 1.36) | 0.0002 |
| **FCMD→CMM** | 1.40 (1.15, 1.70) | 0.0009 |
| **FCMD→Death** | 1.22 (0.98, 1.52) | 0.07 |
| **CMM→Death** | 1.02 (0.61, 1.71) | 0.94 |
| **PM_2.5-10_** |  |  |
| **Baseline→FCMD** | 1.03 (0.96, 1.10) | 0.48 |
| **Baseline→Death** | 1.04 (0.92, 1.18) | 0.49 |
| **FCMD→CMM** | 0.98 (0.77, 1.23) | 0.83 |
| **FCMD→Death** | 0.75 (0.58, 0.98) | 0.03 |
| **CMM→Death** | 1.02 (0.56, 1.87) | 0.94 |
| **PM_10_** |  |  |
| **Baseline→FCMD** | 1.03 (0.99, 1.06) | 0.16 |
| **Baseline→Death** | 1.10 (1.04, 1.17) | <0.0001 |
| **FCMD→CMM** | 1.09 (0.98, 1.23) | 0.12 |
| **FCMD→Death** | 1.10 (0.97, 1.25) | 0.13 |
| **CMM→Death** | 1.10 (0.82, 1.49) | 0.53 |
| **NO_2_** |  |  |
| **Baseline→FCMD** | 1.02 (1.01, 1.02) | <0.0001 |
| **Baseline→Death** | 1.02 (1.01, 1.04) | 0.0007 |
| **FCMD→CMM** | 1.04 (1.01, 1.06) | 0.0032 |
| **FCMD→Death** | 1.04 (1.01, 1.07) | 0.0044 |
| **CMM→Death** | 1.01 (0.95, 1.08) | 0.77 |
| **NO_x_** |  |  |
| **Baseline→FCMD** | 1.01 (1.01, 1.02) | <0.0001 |
| **Baseline→Death** | 1.01 (1.01, 1.02) | <0.0001 |
| **FCMD→CMM** | 1.02 (1.00, 1.03) | 0.008 |
| **FCMD→Death** | 1.02 (1.00, 1.03) | 0.02 |
| **CMD→Death** | 1.01 (0.97, 1.04) | 0.69 |
| **Co-exposure score** |  |  |
| **Baseline→FCMD** | 1.04 (1.03, 1.05) | <0.0001 |
| **Baseline→Death** | 1.05 (1.02, 1.07) | <0.0001 |
| **FCMD→CMM** | 1.07 (1.03, 1.12) | 0.001 |
| **FCMD→Death** | 1.06 (1.01, 1.11) | 0.01 |
| **CMM→Death** | 1.02 (0.91, 1.14) | 0.76 |

a: Models adjusted for age, sex, ethnicity, BMI, years of education, smoking status, moderate alcohol intake, high-level physical activity, total household income, employment status, baseline diet behavior, and levels of low-density lipoprotein, high-density lipoprotein, triglycerides, total cholesterol, systolic blood pressure, and diastolic blood pressure. Estimates of air pollutants were demonstrated per 5-μg/m^3^ increase and estimates of co-exposure score were demonstrated per one SD increase.

Diet behavior was coded as 0-5 based on the sum of the following five items: 1) vegetable intake of at least four tablespoons each day (median); 2) fruit intake of at least three pieces each day (median); 3) fish intake of at least twice each week (median); 4) unprocessed red meat intake of no more than twice each week (median); and 5) processed meat intake of no more than twice each week (median).

**Table S12** Associations of air pollution with the trajectories of cardiometabolic multimorbidity of pattern A using multi-state model additionally adjusted for genetic principal components

|  | **HR (95%CI)** | ***p-*values** |
| --- | --- | --- |
| **PM_2.5_** |  |  |
| **Baseline→FCMD** | 1.31 (1.24, 1.38) | <0.0001 |
| **Baseline→Death** | 1.37 (1.26, 1.50) | <0.0001 |
| **FCMD→CMM** | 1.36 (1.16, 1.60) | <0.0001 |
| **FCMD→Death** | 1.36 (1.14, 1.62) | 0.0007 |
| **CMM→Death** | 0.86 (0.56, 1.33) | 0.50 |
| **PM_2.5-10_** |  |  |
| **Baseline→FCMD** | 1.02 (0.96, 1.08) | 0.62 |
| **Baseline→Death** | 1.02 (0.92, 1.13) | 0.74 |
| **FCMD→CMM** | 0.99 (0.82, 1.21) | 0.95 |
| **FCMD→Death** | 0.97 (0.79, 1.20) | 0.79 |
| **CMM→Death** | 0.93 (0.56, 1.53) | 0.76 |
| **PM_10_** |  |  |
| **Baseline→FCMD** | 1.02 (0.99, 1.05) | 0.12 |
| **Baseline→Death** | 1.11 (1.05, 1.17) | <0.0001 |
| **FCMD→CMM** | 1.09 (0.99, 1.20) | 0.07 |
| **FCMD→Death** | 1.15 (1.04, 1.28) | 0.01 |
| **CMM→Death** | 1.01 (0.79, 1.30) | 0.93 |
| **NO_2_** |  |  |
| **Baseline→FCMD** | 1.02 (1.01, 1.02) | <0.0001 |
| **Baseline→Death** | 1.03 (1.02, 1.04) | <0.0001 |
| **FCMD→CMM** | 1.04 (1.02, 1.06) | <0.0001 |
| **FCMD→Death** | 1.04 (1.02, 1.07) | <0.0001 |
| **CMM→Death** | 1.00 (0.95, 1.05) | 0.98 |
| **NO_x_** |  |  |
| **Baseline→FCMD** | 1.02 (1.01, 1.02) | <0.0001 |
| **Baseline→Death** | 1.02 (1.01, 1.03) | <0.0001 |
| **FCMD→CMM** | 1.02 (1.01, 1.03) | 0.001 |
| **FCMD→Death** | 1.02 (1.01, 1.03) | 0.001 |
| **CMD→Death** | 1.01 (0.99, 1.04) | 0.31 |
| **Co-exposure score^a^** |  |  |
| **Baseline→FCMD** | 1.03 (1.02, 1.04) | <0.0001 |
| **Baseline→Death** | 1.04 (1.03, 1.05) | <0.0001 |
| **FCMD→CMM** | 1.04 (1.02, 1.06) | 0.0002 |
| **FCMD→Death** | 1.05 (1.02, 1.07) | 0.0002 |
| **CMM→Death** | 1.00 (0.95, 1.06) | 0.93 |

### a: Models adjusted for age, sex, BMI, years of education, smoking status, moderate alcohol intake, high-level physical activity, total household income, employment status, and the first ten genetic principal components. Estimates of air pollutants were demonstrated per 5-μg/m^3^ increase and estimates of co-exposure score were demonstrated per one SD increase.

### Table S13 Associations of air pollution with the trajectories of cardiometabolic multimorbidity of pattern A using multi-state model in white participants ^a^

|  | **HR (95%CI)** | ***p-*values** |
| --- | --- | --- |
| **PM_2.5_** |  |  |
| **Baseline→FCMD** | 1.31 (1.25, 1.38) | <0.0001 |
| **Baseline→Death** | 1.39 (1.28, 1.52) | <0.0001 |
| **FCMD→CMM** | 1.39 (1.17, 1.64) | <0.0001 |
| **FCMD→Death** | 1.35 (1.13, 1.61) | <0.0001 |
| **CMM→Death** | 0.86 (0.56, 1.33) | 0.51 |
| **PM_2.5-10_** |  |  |
| **Baseline→FCMD** | 1.02 (0.96, 1.08) | 0.59 |
| **Baseline→Death** | 1.02 (0.92, 1.13) | 0.73 |
| **FCMD→CMM** | 1.01 (0.83, 1.24) | 0.89 |
| **FCMD→Death** | 0.95 (0.77, 1.17) | 0.62 |
| **CMM→Death** | 0.84 (0.50, 1.4) | 0.50 |
| **PM_10_** |  |  |
| **Baseline→FCMD** | 1.02 (0.99, 1.05) | 0.16 |
| **Baseline→Death** | 1.11 (1.05, 1.17) | <0.0001 |
| **FCMD→CMM** | 1.09 (0.99, 1.21) | 0.08 |
| **FCMD→Death** | 1.13 (1.02, 1.25) | 0.02 |
| **CMM→Death** | 1.02 (0.79, 1.32) | 0.88 |
| **NO_2_** |  |  |
| **Baseline→FCMD** | 1.02 (1.01, 1.02) | <0.0001 |
| **Baseline→Death** | 1.03 (1.02, 1.04) | <0.0001 |
| **FCMD→CMM** | 1.04 (1.02, 1.06) | <0.0001 |
| **FCMD→Death** | 1.04 (1.02, 1.07) | <0.0001 |
| **CMM→Death** | 1.00 (0.95, 1.06) | 0.96 |
| **NO_x_** |  |  |
| **Baseline→FCMD** | 1.02 (1.01, 1.02) | <0.0001 |
| **Baseline→Death** | 1.02 (1.01, 1.03) | <0.0001 |
| **FCMD→CMM** | 1.02 (1.01, 1.03) | <0.0001 |
| **FCMD→Death** | 1.02 (1.01, 1.03) | <0.0001 |
| **CMD→Death** | 1.01 (0.98, 1.03) | 0.66 |
| **Co-exposure score^a^** |  |  |
| **Baseline→FCMD** | 1.03 (1.02, 1.04) | <0.0001 |
| **Baseline→Death** | 1.04 (1.03, 1.06) | <0.0001 |
| **FCMD→CMM** | 1.05 (1.02, 1.07) | <0.0001 |
| **FCMD→Death** | 1.05 (1.02, 1.07) | <0.0001 |
| **CMM→Death** | 1.00 (0.94, 1.06) | 0.91 |

a: Models adjusted for age, sex, ethnicity, BMI, years of education, smoking status, moderate alcohol intake, high-level physical activity, total household income, and employment status. Estimates of air pollutants were demonstrated per 5-μg/m^3^ increase and estimates of co-exposure score were demonstrated per one SD increase.

**Table S14** Associations of air pollution with the trajectories of cardiometabolic multimorbidity of pattern A using multi-state model in participants living in the baseline address for more than five years

|  | **HR (95%CI)** | ***p-*values** |
| --- | --- | --- |
| **PM_2.5_** |  |  |
| **Baseline→FCMD** | 1.30 (1.23, 1.37) | <0.0001 |
| **Baseline→Death** | 1.34 (1.21, 1.47) | <0.0001 |
| **FCMD→CMM** | 1.38 (1.16, 1.64) | 0.0002 |
| **FCMD→Death** | 1.32 (1.09, 1.59) | 0.004 |
| **CMM→Death** | 1.00 (0.64, 1.56) | 0.98 |
| **PM_2.5-10_** |  |  |
| **Baseline→FCMD** | 1.02 (0.95, 1.08) | 0.64 |
| **Baseline→Death** | 1.04 (0.93, 1.17) | 0.46 |
| **FCMD→CMM** | 0.96 (0.78, 1.19) | 0.72 |
| **FCMD→Death** | 0.99 (0.79, 1.24) | 0.89 |
| **CMM→Death** | 1.06 (0.63, 1.79) | 0.81 |
| **PM_10_** |  |  |
| **Baseline→FCMD** | 1.02 (0.99, 1.05) | 0.19 |
| **Baseline→Death** | 1.11 (1.05, 1.17) | 0.0001 |
| **FCMD→CMM** | 1.08 (0.98, 1.20) | 0.13 |
| **FCMD→Death** | 1.15 (1.03, 1.29) | 0.01 |
| **CMM→Death** | 1.08 (0.83, 1.41) | 0.55 |
| **NO_2_** |  |  |
| **Baseline→FCMD** | 1.02 (1.01, 1.02) | <0.0001 |
| **Baseline→Death** | 1.03 (1.02, 1.04) | <0.0001 |
| **FCMD→CMM** | 1.04 (1.02, 1.06) | <0.0001 |
| **FCMD→Death** | 1.05 (1.02, 1.07) | <0.0001 |
| **CMM→Death** | 1.01 (0.95, 1.06) | 0.85 |
| **NO_x_** |  |  |
| **Baseline→FCMD** | 1.02 (1.01, 1.02) | <0.0001 |
| **Baseline→Death** | 1.02 (1.01, 1.03) | <0.0001 |
| **FCMD→CMM** | 1.02 (1.01, 1.03) | 0.0007 |
| **FCMD→Death** | 1.02 (1.01, 1.03) | 0.004 |
| **CMD→Death** | 1.02 (0.99, 1.05) | 0.13 |
| **Co-exposure score^a^** |  |  |
| **Baseline→FCMD** | 1.03 (1.02, 1.04) | <0.0001 |
| **Baseline→Death** | 1.04 (1.03, 1.05) | <0.0001 |
| **FCMD→CMM** | 1.04 (1.02, 1.07) | 0.0001 |
| **FCMD→Death** | 1.04 (1.02, 1.07) | 0.001 |
| **CMM→Death** | 1.02 (0.96, 1.08) | 0.52 |

a: Models adjusted for age, sex, ethnicity, BMI, years of education, smoking status, moderate alcohol intake, high-level physical activity, total household income, and employment status. Estimates of air pollutants were demonstrated per 5-μg/m^3^ increase and estimates of co-exposure score were demonstrated per one SD increase.

### Table S15 Associations of air pollution with the trajectories of cardiometabolic multimorbidity of pattern A using multi-state model by the age of the onset of first cardiometabolic disease ^a^

|  | **Age of FCMD onset** | | | | |  |
| --- | --- | --- | --- | --- | --- | --- |
|  | **<65 years** | | **≥65 years** | | | ***p*-values of interaction** |
|  | **HR (95%CI)** | ***p-*values** | **HR (95%CI)** | ***p-*values** | |  |
| **PM_2.5_** | 1.48 (1.15, 1.92) | <0.0001 | 1.25 (1.01, 1.55) | | 0.04 | 0.382 |
| **PM_2.5-10_** | 1.06 (0.78, 1.45) | 0.69 | 1.05 (0.81, 1.35) | | 0.71 | 0.979 |
| **PM_10_** | 1.21 (1.04, 1.40) | 0.01 | 1.04 (0.92, 1.18) | | 0.53 | 0.228 |
| **NO_2_** | 1.02 (1.01, 1.04) | <0.0001 | 1.02 (1.00, 1.03) | | 0.03 | 0.261 |
| **NO_x_** | 1.06 (1.02, 1.09) | <0.0001 | 1.03 (1.00, 1.05) | | 0.05 | 0.454 |
| **Co-exposure score** | 1.09 (1.04, 1.16) | <0.0001 | 1.05 (1.00, 1.10) | | 0.03 | 0.318 |

a: Models adjusted for age, sex, ethnicity, BMI, years of education, smoking status, moderate alcohol intake, high-level physical activity, total household income, and employment status. Estimates of air pollutants were demonstrated per 5-μg/m^3^ increase and estimates of co-exposure score were demonstrated per one SD increase. Interaction *p*-values were retrieved from models with the interaction term of age group and air pollution.
